# Unidirectional and Bidirectional Causation between Smoking and Blood DNA Methylation: Evidence from Twin-based Mendelian Randomisation

**DOI:** 10.1101/2024.06.19.24309184

**Authors:** Madhurbain Singh, Conor V. Dolan, Dana M. Lapato, Jouke-Jan Hottenga, René Pool, Brad Verhulst, Dorret I. Boomsma, Charles E. Breeze, Eco J. C. de Geus, Gibran Hemani, Josine L. Min, Roseann E. Peterson, Hermine H. M. Maes, Jenny van Dongen, Michael C. Neale

## Abstract

Cigarette smoking is associated with numerous differentially-methylated genomic loci in multiple human tissues. These associations are often assumed to reflect the causal effects of smoking on DNA methylation (DNAm), which may underpin some of the adverse health sequelae of smoking. However, prior causal analyses with Mendelian Randomisation (MR) have found limited support for such effects. Here, we apply an integrated approach combining MR with twin causal models to examine causality between smoking and blood DNAm in the Netherlands Twin Register (N=2577). Analyses revealed potential causal effects of current smoking on DNAm at >500 sites in/near genes enriched for functional pathways relevant to known biological effects of smoking (e.g., hemopoiesis, cell- and neuro-development, and immune regulation). Notably, we also found evidence of reverse and bidirectional causation at several DNAm sites, suggesting that variation in DNAm at these sites may influence smoking liability. Seventeen of the loci with putative effects of DNAm on smoking showed highly specific enrichment for gene-regulatory functional elements in the brain, while the top three sites annotated to genes involved in G protein-coupled receptor signalling and innate immune response. These novel findings are partly attributable to the analyses of *current* smoking in twin models, rather than *lifetime* smoking typically examined in MR studies, as well as the increased statistical power achieved using multiallelic/polygenic scores as instrumental variables while controlling for potential horizontal pleiotropy. This study highlights the value of twin studies with genotypic and DNAm data for investigating causal relationships of DNAm with health and disease.

## Introduction

Epigenome-wide association studies (EWASs) identify variation in DNA methylation (DNAm) associated with complex human traits and diseases [1]. Arguably, the most successful EWASs have been studies of cigarette smoking. A large-scale EWAS meta-analysis of current versus never smoking revealed significant DNAm differences at 18,760 CpG (*Cytosine-phosphate-Guanine*) sites in peripheral blood cells [2]. DNAm differences between former- and never-smoking individuals were diminished but remained significant at 2,568 sites. Genes annotated to the differentially methylated CpGs have been implicated in genome-wide association studies (GWAS) of numerous smoking-associated traits, including cancers, lung functions, cardiovascular disorders, inflammatory disorders, and schizophrenia [2].

As standard cross-sectional EWAS in unrelated individuals cannot differentiate between causation and confounding [3], different etiological mechanisms may underlie the associations between cigarette smoking and DNAm. These associations are typically interpreted as the causal *effects* of smoking exposure on DNAm. However, some smoking-associated CpGs may have reverse or bidirectional causal links with smoking, i.e., DNAm may reciprocally affect the development and maintenance of smoking behaviours [4]. Moreover, associations between smoking and DNAm may be attributable to confounders, such as schizophrenia [5], alcohol [6] and cannabis use [7] and body mass index [8].

Mendelian Randomisation (MR) analyses use genetic variants as instrumental variables (IVs) to estimate causal effects [3,9]. MR analyses have identified the effects of lifetime (current or former) smoking on blood DNAm at only 11 CpGs [10], with reverse effects of blood DNAm at nine sites [11]. Causal inference in MR is based on the assumption that the genetic variants associated with the exposure influence the outcome exclusively through the exposure. Specifically, genetic IVs for smoking may show vertical, but not horizontal, pleiotropy with DNAm. To minimise the risk of horizontal pleiotropy, MR analyses require carefully selected single-nucleotide polymorphisms (SNPs), including using genetic colocalisation to filter out SNPs showing horizontal pleiotropy due to linkage disequilibrium (LD). Since SNPs usually have small effect sizes, traditional MR approaches may have limited power to detect causality and may be subject to weak-instrument bias [12]. Furthermore, causal inference in standard, summary-statistics-based MR analyses typically applies to the GWAS phenotype of *lifetime* smoking. However, as most smoking-associated DNAm changes exhibit substantial reversibility upon smoking cessation [2,13], it is important to examine the causal relationships of *current* smoking specifically.

Recent methodological developments integrate the principles of MR with the twin-based *Direction of Causation* (DOC) model [14], giving rise to the unidirectional *MR-DoC1* [15] and the bidirectional *MR-DoC2* models [16]. MR-DoC1 allows one to estimate and account for horizontal pleiotropy, while MR-DoC2 accommodates pleiotropy arising from LD. Thus, these models enable using polygenic risk scores (PRS) as IVs, increasing the statistical power to estimate causal effects and curtailing weak-instrument bias, relative to MR methods using individual SNPs as IVs. Incorporating MR with family data also helps to resolve additional assumptions of standard MR, such as random mating and no dyadic effects [15,17]. Moreover, by using participant-level information, these models estimate causal effects between the phenotypes measured in the twins, allowing separate causal models for current and former smoking.

The present study used MR-DoC models to examine bidirectional causal effects between cigarette smoking and peripheral blood DNAm in a population-based cohort of European ancestry adult twins from the Netherlands Twin Register (NTR) [18,19]. The target sample included 2,577 individuals from 1,459 twin pairs with both genotypic and DNAm data, and self-reported smoking status at the time of blood draw. Across 16,940 smoking-related CpGs, we fitted separate models for current (versus never) and former (versus never) smoking. We obtained a set of three causal estimates in each direction (*Smoking → DNAm*, *DNAm → Smoking*): the estimates from bidirectional MR-DoC2, and two different model specifications of unidirectional MR-DoC1 (**Figure 1**). We triangulated evidence across the three models based on the statistical significance and consistency of the causal estimates. The results indicated much more widespread putative causal influences of current smoking on DNAm than *vice versa*. Follow-up enrichment analyses highlighted biological processes and tissues relevant to the CpGs with potential effects in either direction of causation.

**Figure 1.**
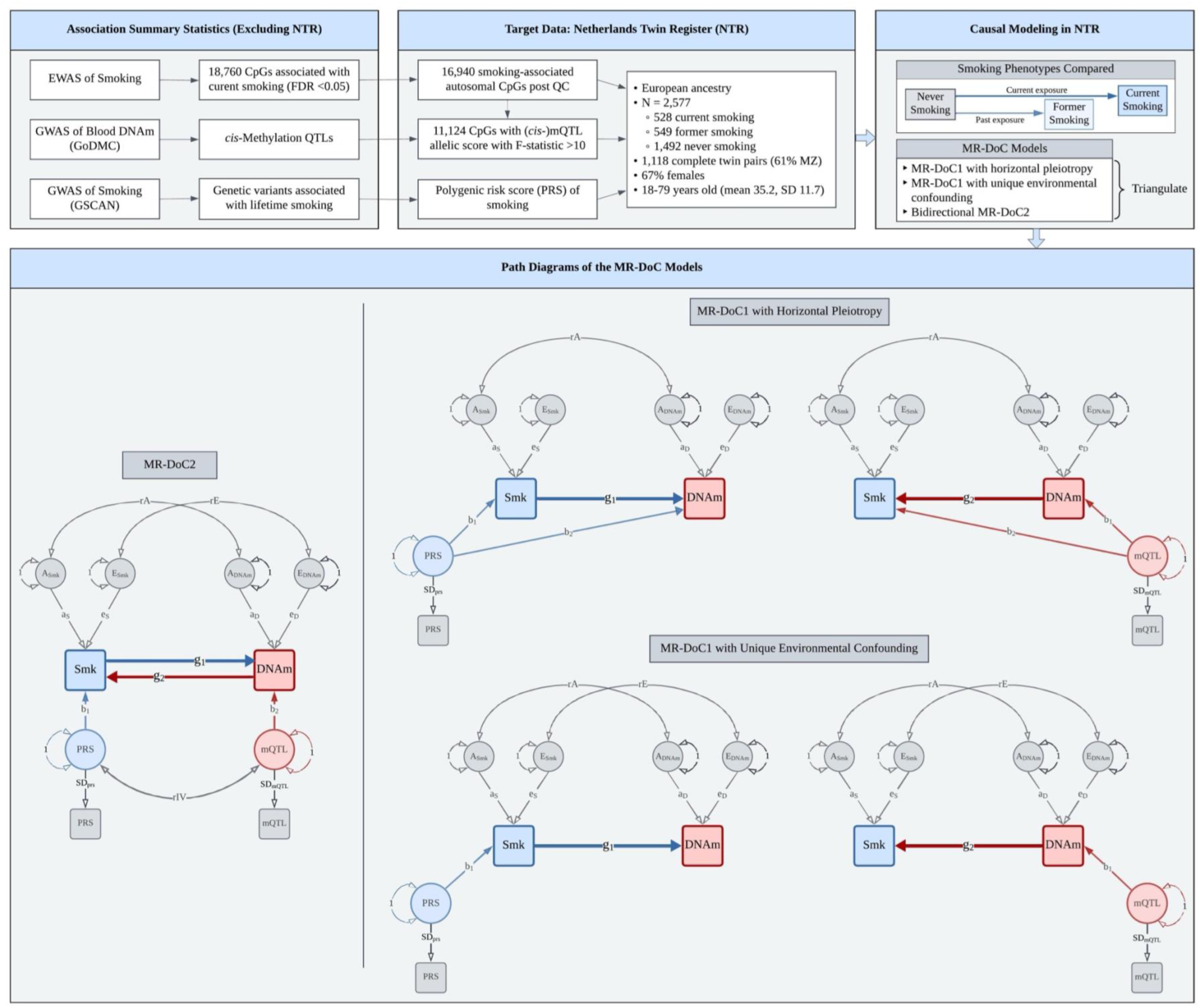
Study Design. Overview of the data and MR-DoC models used to examine the causality between cigarette smoking and blood DNA methylation (DNAm) in the Netherlands Twin Register. The models were fitted separately for current (versus never) and former (versus never) smoking. Applying the five MR-DoC models shown in the path diagrams, we obtained a set of three causal estimates in each direction of causation: Smoking (Smk) → DNAm (the blue paths labelled **g_1_**) and DNAm → Smoking (the red paths labelled **g_2_**). In each MR-DOC model, the residual variance of each phenotype (smoking status liability and DNAm levels) is decomposed into latent additive genetic (A) and unique environmental (E) factors. The correlation between the latent A factors of smoking and DNAm (rA) represents confounding due to additive genetic factors, while that between the latent E factors (rE) represents confounding due to unique environmental factors. Note that these models did not include shared environmental (C) variance components, as the AE model was found to be the most parsimonious in univariate twin models (see **Supplementary Methods**). Note. For better readability, the path diagrams show only the within-individual part of the models fitted to data from twin pairs. The squares/rectangles indicate observed variables, the circles indicate latent (unobserved) variables, the single-headed arrows indicate regression paths, and the double-headed curved arrows indicate (co-)variances.

## Methods

### Study Sample

We analysed data from 706 monozygotic (MZ) twin pairs, 412 dizygotic (DZ) twin pairs, and 341 individuals without their co-twin. The participants, 1,730 (67%) females and 847 (33%) males, were aged 18–79 (mean 35.2; S.D. 11.7 years) at the time of blood draw. Sample and variant quality control (QC) of genotypic data, imputation, genetic principal component analysis (PCA), and ancestry-outlier pruning have been described previously [20], and reviewed in **Supplementary Methods**. Since the summary statistics of methylation quantitative trait loci (mQTLs) were available for European ancestry only [21], we excluded 109 participants identified as European-ancestry outliers to avoid bias due to ancestry mismatch.

The NTR is approved by the Central Ethics Committee on Research Involving Human Subjects of the VU University Medical Centre, Amsterdam, an Institutional Review Board certified by the U.S. Office of Human Research Protections (IRB number IRB00002991 under Federal-wide Assurance-FWA00017598; IRB/institute codes, NTR 98-222, 2003-180, 2008-244). All participants provided written informed consent before data collection.

### Peripheral Blood DNA Methylation and Cell Counts

Epigenome-wide DNAm in peripheral whole blood was measured with the Infinium HumanMethylation450 BeadChip Kit (“Illumina 450k” microarray), following manufacturer’s protocol [22]. DNAm data QC and normalisation were performed using a custom pipeline developed by the BIOS (Biobank-based Integrative Omics Study) Consortium [23] (**Supplementary Methods**). In the current analyses, only autosomal probes were included, yielding 411,169 CpGs that passed QC, of which 16,940 sites were associated with current smoking (FDR <0.05) in a previous independent EWAS [2] (hereafter called the “smoking-associated CpGs”). These CpGs were analysed in the MR-DoC1 models for *Current Smoking → DNAm* (**Figure 2**). Likewise, 2,330 autosomal, post-QC CpGs, previously associated with former smoking [2] (hereafter called the “former-smoking-associated CpGs”), were analysed in the MR-DoC1 models for *Former Smoking → DNAm*. Differential white blood cell counts were also measured in the blood samples [23].

**Figure 2.**
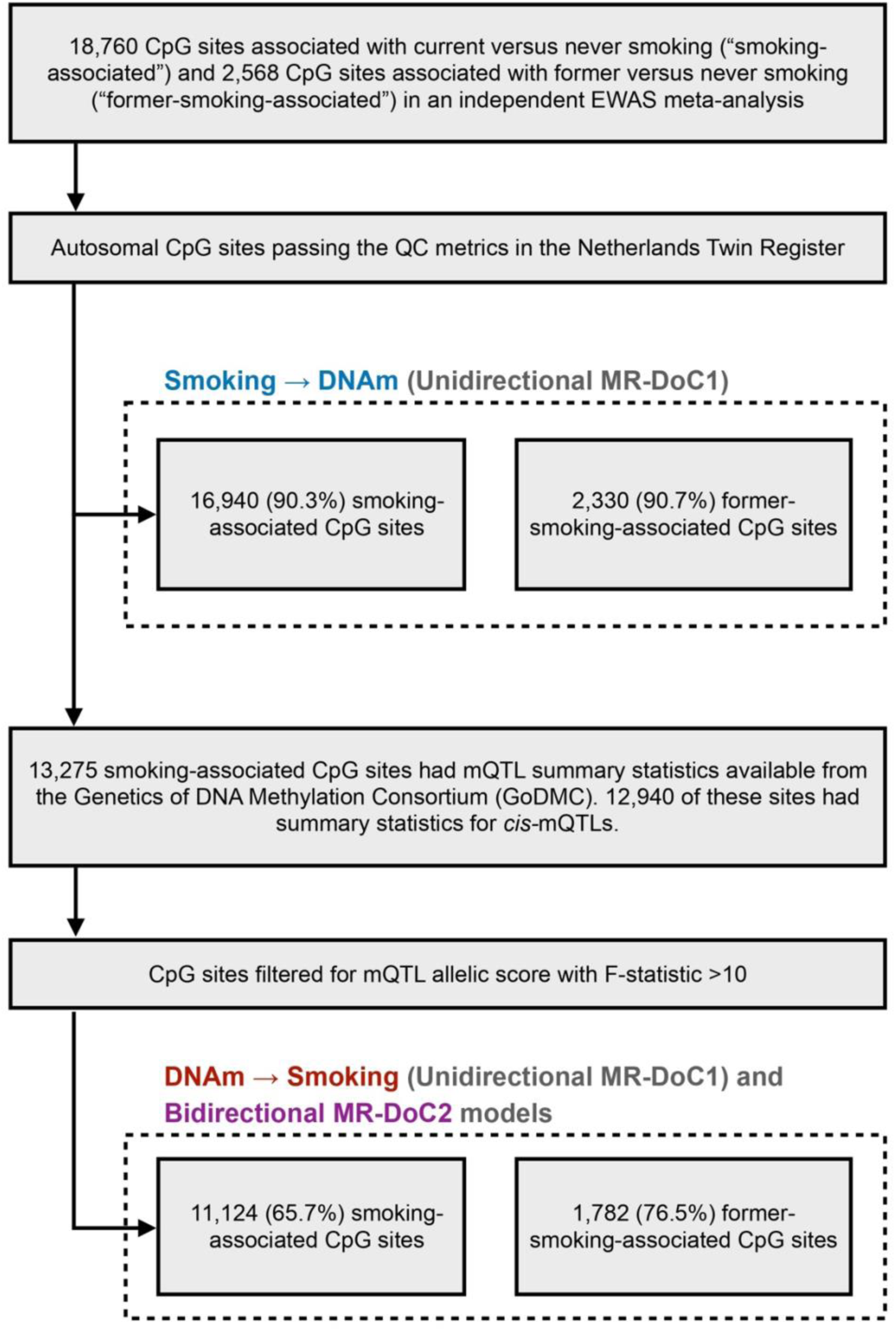
Selection of CpGs tested in each MR-DoC model. Previous independent EWAS meta-analysis of cigarette smoking [2] examined DNA methylation (DNAm) at CpGs from the Illumina HumanMethylation450 BeadChip array [22], which was also used to measure DNAm in the NTR biobank. In the unidirectional MR-DoC1 models for Smoking → DNAm, we included autosomal CpGs associated with smoking in the EWAS meta-analysis that also passed the QC metrics in NTR. The MR-DoC1 models for DNAm → Smoking and the bidirectional MR-DoC2 models were restricted to a subset of these sites having cis-mQTL summary statistics from the GoDMC [21] and a resulting mQTL allelic score with F-statistic >10.

The normalised β-values of DNAm at each CpG were residualised by regressing out age, sex (genotypically inferred biological sex, matched with self-reported sex), measured white blood cell percentages (neutrophils, monocytes, and eosinophils), HM450k array row, and bisulfite sample plate [24]. The residuals were standardised (mean = 0, S.D. = 1). As in the previous NTR work [24], we excluded lymphocyte percentage as a covariate, given its multicollinearity with neutrophil percentage. We excluded basophil percentage because of its low variance.

### Cigarette Smoking

Self-reported cigarette smoking status was recorded during blood sample collection in 2004-2008 and 2010-2011 (**Supplementary Methods**), with the question, “Do you smoke?” with three response options: “No, I never smoked” (N = 1,492), “No, but I did in the past” (N = 549), and “Yes” (N = 528). The responses were checked for consistency with data from the longitudinal NTR surveys and plasma cotinine levels (**Supplementary Methods**).

### Instrumental Variables

#### mQTL allelic scores

We used a weighted sum of DNAm-increasing alleles at *cis-*mQTLs (“mQTL allelic score”) as the IV for DNAm, computed using clumping and thresholding in *PLINK1.9* [25] (**Supplementary Methods**). Of the 16,940 smoking-associated CpGs, 12,940 had summary statistics for *cis-*mQTLs available from the Genetics of DNA Methylation Consortium (GoDMC; excluding NTR) [21] (**Figure 2**). We used only *cis-*mQTLs, i.e., SNPs within 1Mb of the CpG, given that SNPs located close to the CpG are likely to be associated with smoking *via* DNAm. To further guard against potential horizontal pleiotropy with smoking, we relied on the consistency of the causal estimates in MR-DoC models accommodating horizontal pleiotropy. To reduce the risk of weak-instrument bias, we restricted the MR-DoC1 models for *DNAm → Current Smoking* and the bidirectional MR-DoC2 models to 11,124 (65.7%) CpGs having an mQTL allelic score with F-statistic >10 (**Figure 2**). The included mQTL allelic scores had an incremental R^2^ for the respective CpG site ranging from 0.43% to 76.95% (mean 9.04%, S.D. = 10.94%). Similarly, a subset of 1,782 (76.5%) former-smoking-associated CpGs had mQTL allelic scores with F-statistic >10 and were examined in the MR-DoC1 models for *DNAm → Former Smoking* and the bidirectional model (MR-DoC2).

#### PRS of Regular Smoking Initiation

We used a PRS of lifetime regular-smoking initiation as the IV for smoking status, computed using *LDpred v0.9* [26] with European-ancestry GWAS summary statistics [27] (**Supplementary Methods**). This PRS had an incremental liability-scale R^2^ of 5.07% (F-statistic = 73.2) for current versus never smoking, and 2.02% (F-statistic = 28.8) for former versus never smoking. The smoking phenotypes in MR-DoC models differed from the GWAS phenotype (smoking initiation = current/former versus never smoking). However, in these causal models, the strength of the IV, the extent of horizontal pleiotropy with DNAm, and the estimated causal effects on DNAm apply to the smoking phenotype operationalised in the target data.

We residualised the smoking PRS and all mQTL allelic scores for the genotyping platform and the first ten genetic PCs, and standardised the residuals (mean = 0, S.D. = 1).

### MR-DoC Models

Causal inference in twin data leverages the cross-twin cross-trait correlations to estimate the direction and magnitude of potential causal effects between traits [14]. On the other hand, MR analyses rely on the assumptions that the IV is (1) associated with the exposure (“relevance”), (2) not correlated with any omitted confounding variables (“exchangeability”), and (3) independent of the outcome, given the exposure (“exclusion restriction”) [3,28]. Here, we used the criterion of F-statistic >10 to identify “relevant” IVs. Further, genetic variants are assumed to satisfy the “exchangeability” assumption, given Mendel’s laws of random segregation and independent assortment. The “exclusion restriction” assumption for a genetic IV implies no horizontal pleiotropy with the outcome. Here, we applied different MR-DoC models (**Figure 1**) to account for possible horizontal pleiotropy. MR-DoC1 accommodates horizontal pleiotropy under the assumption of no confounding due to unique environmental factors. The alternative specification of MR-DoC1 accommodates unique environmental confounding (parameter “rE” in **Figure 1**), given the assumption of no horizontal pleiotropy [15]. In both cases, the model includes confounding due to genetic and shared environmental influences on the exposure and the outcome. In MR-DoC2 models, we estimated bidirectional causal effects by including the smoking PRS and the mQTL allelic score, allowing the two IVs to covary [16]. Beyond the causal effects between smoking and DNAm, the covariance between the PRS and the mQTL allelic score may arise from several sources, including shared pleiotropic SNPs, LD between the constituent SNPs, and population structure. By accommodating these sources of covariance, MR-DoC2 may help reduce potential biases in the causal estimates.

The MR-DoC models were fitted in the *OpenMx* package (v2.21.8) [29] in R (v4.3.2), using the code from the original publications [15,16] (**Supplementary Methods**). Binary smoking status was examined in the liability threshold model [30], so the causal estimate is interpreted as the effect of the underlying smoking *liability* rather than smoking *exposure*. Age and sex were included in the model as covariates of smoking status. For each set of causal estimates across CpGs (**Figure 1**), we calculated the Bayesian genomic inflation factor (λ) using the R package *bacon* [31], made QQ plots using the R package *GWASTools* [32], and applied Benjamini-Hochberg FDR correction [33] to the p-values.

### Functional Enrichment Analyses

We used *Metascape* (v3.5.20240101) [34] to perform gene-set annotation and functional enrichment analyses of the CpGs with potential causal effects in either direction (**Supplementary Methods**). The input list of gene IDs was selected based on proximity to the CpGs with consistent and nominally significant (p<0.05) estimates in all three models.

Furthermore, to explore the tissue-specific functional relevance of the implicated CpGs, we performed *eFORGE 2.0* (experimentally derived Functional element Overlap analysis of ReGions from EWAS) analyses [35–37]. We examined the overlap between the implicated CpGs and multiple comprehensive reference sets of tissue-/cell type-specific gene regulatory genomic and epigenomic features, including chromatin states, histone marks, and DNase-I hotspots (**Supplementary Methods**).

## Results

### Exemplar: Putative causality between current smoking and *AHRR* DNAm

To illustrate the three MR-DoC models, we first present the results for two CpGs (cg23916896 and cg05575921) in the Aryl-Hydrocarbon Receptor Repressor (*AHRR*) gene, with well-established DNAm associations with cigarette smoking [2].

For probe cg23916896 (**Supplement Figure S1A**), the mQTL allelic score had an incremental R^2^ of 8.03% (F-statistic = 156.4). The MR-DoC models indicated that higher liability for current smoking likely causes hypomethylation of cg23916896, with statistically significant (FDR <0.05), consistently negative causal estimates in all three models. The reverse effect of cg23916896 methylation on the liability for current smoking had consistent negative estimates. However, the estimates were significant at FDR <0.05 in MR-DoC1 with horizontal pleiotropy, but only nominally significant (p <0.05) in the other two models. Taken together, these results provide robust evidence for current smoking’s causal effects on cg23916896 methylation, with suggestive evidence for reverse causation. Previous MR studies have not examined this CpG site, as these studies focused on a few pre-selected sites [10,11]. Our results indicate a potential bidirectional causal relationship between cigarette smoking and cg23916896, i.e., smoking-induced hypomethylation at this locus may reciprocally increase smoking liability.

In comparison, probe cg05575921 had an mQTL allelic score with an incremental R^2^ of 1.74% (F-statistic = 31.6). Similar to cg23916896, the effect of current smoking liability on cg05575921 methylation was consistently negative, with FDR <0.05 in all three models (**Supplement Figure S1B**). This aligns with the previously reported negative, albeit non-significant, effect of *lifetime* smoking [10]. For the reverse effect of cg05575921 methylation on smoking liability, the estimates were negative in all three models, though statistically significant only in MR-DoC1 with horizontal pleiotropy. Notably, the estimates for cg05575921 are comparable to those for cg23916896, but have larger standard errors, likely due to the weaker IV of the former (mQTL allelic score). This variability in the precision of the causal estimates underscores the differences in the strength of the IV across CpGs and, consequently, the power to estimate their causal effect on smoking.

### Evidence of more widespread effects of current smoking on DNAm than *vice versa*

We used genomic inflation factor, λ, to evaluate potential widespread, small causal effects of current smoking on DNAm. Across the smoking-associated CpGs, MR-DoC1 including horizontal pleiotropy (rE = 0) had λ = 1.44, while MR-DoC1 with unique environmental confounding, but no horizontal pleiotropy, showed λ = 1.20. For comparison, fitting similar models epigenome-wide showed less inflation (λ = 0.98 and λ = 1.09, respectively), suggesting enrichment of low p-values among the smoking-associated CpGs, as also reflected in the QQ plots (**Supplementary Figures S2-S3**). The epigenome-wide inflation is consistent with that for cigarettes per day (λ >1.1), as seen in prior two-sample MR analyses [21]. In MR-DoC2 models, the estimated reverse effects of DNAm on current smoking showed little inflation (λ = 1.01) compared to current smoking’s effects on DNAm in the same model (λ = 1.20; **Supplementary Figures S4-S5**). These findings suggest that the causal influences of current smoking on DNAm contribute partly to the previously reported EWAS hits. However, for the reverse effects of DNAm on current smoking, the absence of λ inflation does not preclude potential localised small effects, albeit at fewer CpGs.

There was considerable variability in the number of CpGs with statistically significant causal estimates across models (**Figure 3; top panel**), with a relatively higher number of significant estimates in MR-DoC1 with horizontal pleiotropy, likely due to its higher power [38]. Looking at the intersection of significant *Current Smoking → DNAm* estimates across models, 259 CpGs showed FDR <0.05 in at least two models, while 64 sites showed FDR <0.05 in all three models. These 64 sites also showed a consistent direction of effect in all models (**Supplementary Figure S6, Table S1**). Thus, we considered these 64 CpGs to exhibit robust evidence for current smoking’s effects on DNAm, including hypomethylation of 59 sites and hypermethylation of the other five (**Figure 3; bottom panel**). These CpGs annotate to several top genes implicated in prior EWAS of smoking [2], including hypomethylation of CpGs in/near *AHRR*, *ALPPL2*, *CNTNAP2*, and *PARD3* and hypermethylation of CpGs in *MYO1G*. Only one of these 64 CpGs lies within the major histocompatibility complex (MHC) region: cg06126421 (near *HLA-DRB5*). Due to its complex LD structure, the causal estimates of the sites in the MHC region should be interpreted with caution.

**Figure 3.**
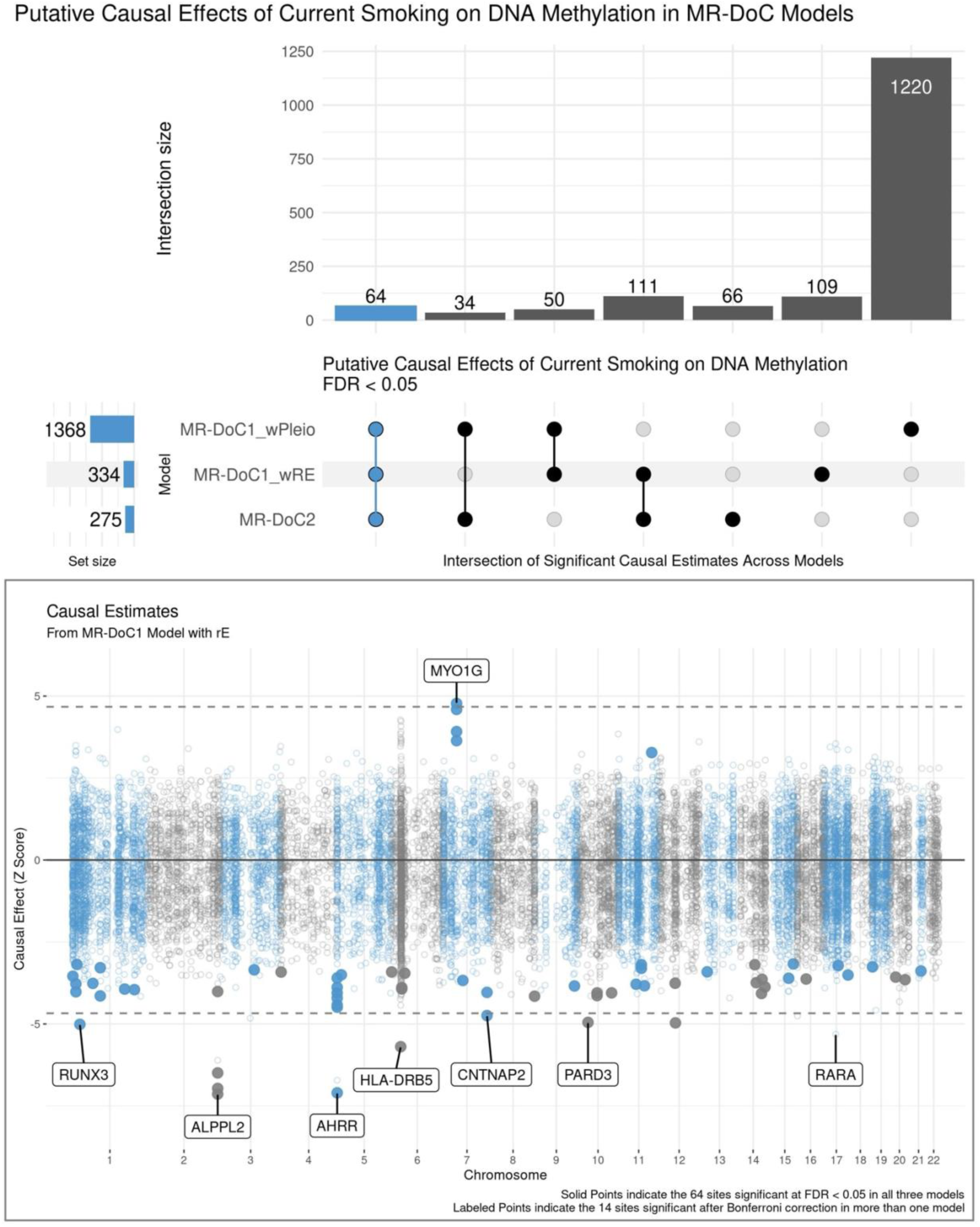
Putative Causal Effects of Current Smoking on Blood DNA Methylation in MR-DoC Models. The top panel shows an UpSet plot of the intersection of CpGs with statistically significant (FDR <0.05) estimates of Current Smoking → DNAm in the three MR-DoC models. The matrix consists of the models along the three rows and their intersections along the columns. The horizontal bars on the left represent the number of CpGs with significant (FDR <0.05) causal estimates in each model. The vertical bars represent the number of CpGs belonging to the respective intersection in the matrix. A similar UpSet plot with Bonferroni correction is shown in **Supplementary Figure S7**. The bottom panel shows a Miami plot of the Current Smoking → DNAm causal estimates across 16,940 smoking-associated CpGs. The X-axis shows the genomic positions of the CpGs aligned to Genome Reference Consortium Human Build 37 (GRCh37). The Y-axis shows the Z-statistic of the estimated effect of the liability for current (versus never) smoking on (residualised and standardised) DNA methylation b-values in the MR-DoC1 model with unique environmental confounding (rE). The solid points indicate the 64 sites with significant causal estimates (FDR <0.05) in all three models (i.e., the blue vertical bar in the UpSet plot). The 14 CpGs with causal estimates significant after Bonferroni correction in more than one model are labelled by their respective nearest gene. Note. The data underlying these plots are in **Supplementary Table S1**.

For *DNAm → Current Smoking*, 44 CpGs showed FDR <0.05 in at least two models, but only three CpGs had FDR <0.05 in all models (**Figure 4B**). The three CpGs also had consistent, positive estimates across models, suggesting that hypermethylation of CpGs in *GNG7*, *RGS3*, and *SLC15A4* genes may increase smoking liability (**Figure 4A**). None of these sites has been previously reported to influence smoking liability [11].

**Figure 4.**
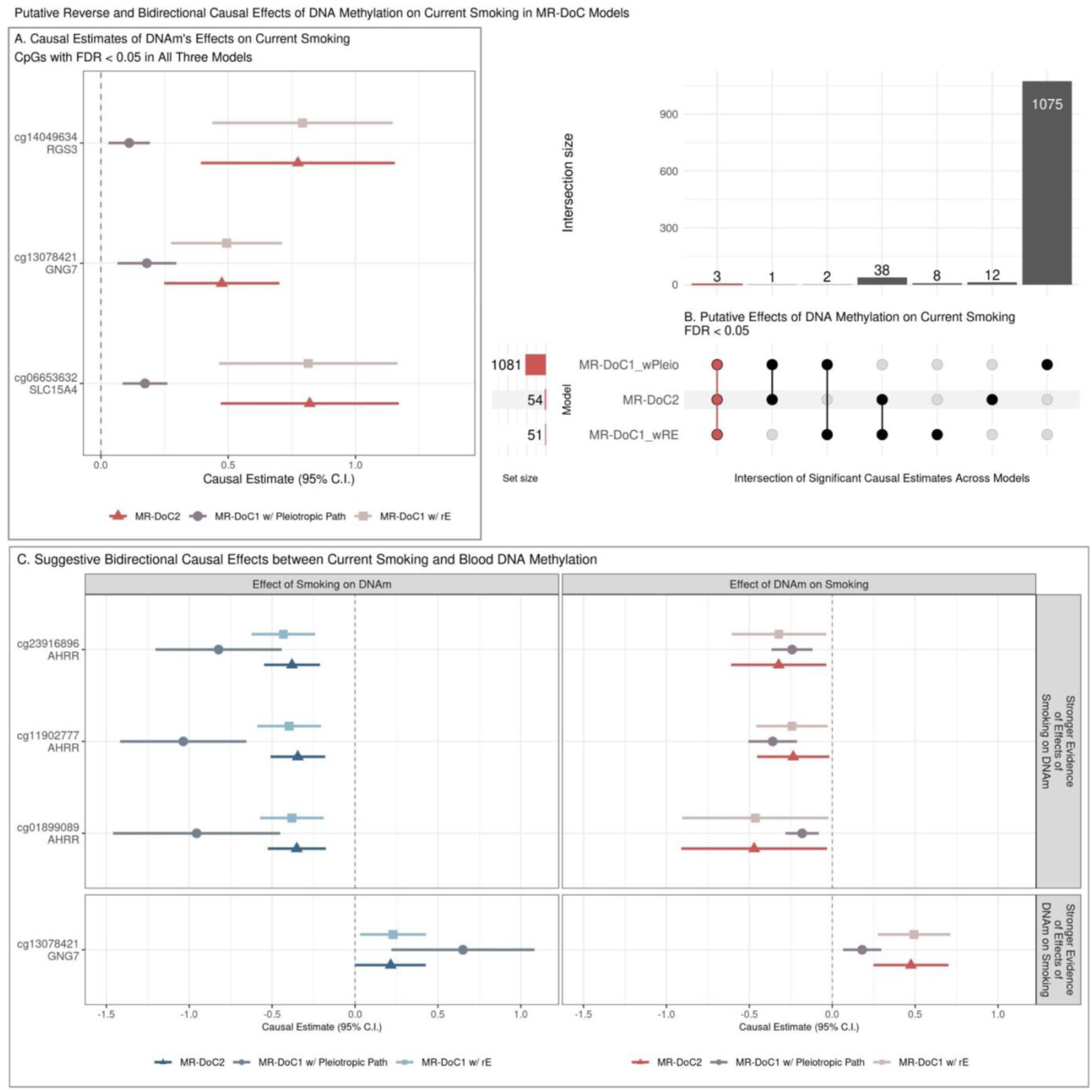
Potential reverse and bidirectional effects of blood DNA methylation on current smoking. **(A.)** Estimates and Wald-type 95% confidence intervals of DNAm → Current Smoking causal effects in each of the three MR-DoC models: bidirectional MR-DoC2, MR-DoC1 with horizontal pleiotropic path, and MR-DoC1 with unique environmental confounding (rE). **(B.)** An UpSet plot of the intersection of CpGs with statistically significant (FDR <0.05) estimates of DNAm → Current Smoking in each of the three MR-DoC models. The matrix consists of the models along the three rows and their intersections along the columns. The horizontal bars on the left represent the number of CpGs with significant (FDR <0.05) causal estimates in each model. The vertical bars represent the number of CpGs belonging to the respective intersection in the matrix. A similar UpSet plot with Bonferroni correction is shown in **Supplementary Figure S8** for comparison. **(C.)** Estimates and Wald-type 95% confidence intervals of bidirectional causal effects between current smoking and DNA methylation in the three MR-DoC models. In panels A and C, the Y-axis labels indicate the CpG probe IDs and the respective genes in which the CpGs are located. Note. The numerical data underlying these plots are in **Supplementary Tables S1-S4**.

### Suggestive Evidence of Bidirectional Effects

Of the 64 sites with robust evidence of *Current Smoking → DNAm* effects, three sites also had consistently negative, nominally significant (p <0.05) estimates of reverse *DNAm → Current Smoking* effects (**Figure 4C**). The three CpGs (cg23916896, cg11902777, cg01899089) are located in the *AHRR* gene, suggesting that current smoking may cause hypomethylation of CpGs in *AHRR*, which may reciprocally increase smoking liability. Among the CpGs with robust evidence of DNAm effects on current smoking, cg13078421 (*GNG7*) also showed consistently positive, nominally significant estimates of current smoking’s effects on DNAm. Thus, *GNG7* hypermethylation increases smoking liability, with a potential reverse effect of current smoking on *GNG7* methylation. Additionally, 15 CpGs had consistent, nominally significant bidirectional causal estimates in all three models, though not significant after FDR correction in either direction (**Supplementary Figure S9**).

### DNAm loci potentially influenced by smoking are enriched for biological processes relevant to smoking’s adverse health outcomes

In follow-up functional enrichment analyses, we identified 525 CpGs with potential *Current Smoking → DNAm* effects (excluding 21 sites in the MHC region), based on consistent, nominally significant estimates in all models (**Supplementary Table S1**). The mapped genes showed extensive significant enrichment (FDR <0.05) for ontology clusters, including hemopoiesis, cell morphogenesis, inflammatory response, regulation of cell differentiation, and regulation of nervous system development, underscoring DNAm’s potential role in the adverse health sequelae of smoking (**Supplementary Figures S10-S12**; **Tables S5-S6**). In the *eFORGE* analyses, these sites were significantly enriched (FDR <0.05) for overlap with a wide range of gene regulatory elements in most of the tissue/cell types in reference datasets, suggesting pervasive functional consequences of smoking’s effects on DNAm (**Supplementary Figures S13-S15**; **Tables S7-S9**).

### CpGs with consistent effects on current smoking show enrichment for brain-related gene regulatory elements

We identified 64 CpGs with potential *DNAm → Current Smoking* effects (none in the MHC region), as indicated by consistent, nominally significant estimates across models (**Supplementary Figure S16**). Gene-set enrichment analyses revealed no significant functional enrichment (FDR <0.05), likely due to too few loci (**Supplementary Figures S17-S18**; **Tables S10-S11**). However, the *eFORGE* analyses, which use precise chromatin-based information for each CpG, showed significant enrichment (FDR <0.05) for overlap with enhancers in the brain, blood (primary B cells, hematopoietic stem cells), lung, and mesodermal embryonic stem cells (**Supplementary Figures S19-S21**; **Tables S12-S14**). These CpGs also showed significant enrichment for histone marks in multiple tissues/cell types (including the brain, blood, and lung), though the overlap with DNase-I hotspots was not significantly enriched. The tissues/cell types predicted to be relevant for DNAm’s effects on smoking liability may be prioritised for follow-up functional studies.

To further gauge the tissue-specificity of *eFORGE* enrichment, we performed iterative follow-up analyses with the CpGs overlapping with tissue/cell types of interest (**Supplementary Figures S22-S24; Tables S15-S17**). These analyses elucidated a subset of 17 CpGs with significant and highly specific enrichment for enhancers and histone marks (H3K4me1 and H3K4me4) in the brain (**Figure 5**), along with weaker enrichment for H3K4me1 in the adrenal gland and thymus. Ten of the 17 sites also overlapped with DNase-I hotspots in the brain, though the enrichment was not statistically significant (FDR = 0.08) (**Supplementary Figure S25, Table S20**). The causal estimates and mapped genes of these 17 CpGs are shown in **Supplementary Figure S26**. Four of these CpGs also had consistent estimates of current smoking’s effects on DNAm (identified by the column “g1_nominal” in **Supplementary Table S4**): cg25612391 (*SLC25A42*), cg05424060 (*GNAI1*), cg10590964 (near *KIAA2012*), and cg05877788 (*TP53I13*). Furthermore, prior pre-clinical and clinical studies have implicated 14 of the 17 mapped genes, including three with potential bidirectional effects, in behavioural or neurological traits, including alcohol dependence (*OSBPL5*) [39], cocaine use (*SLCO5A1*) [40], anxiety (*CCDC92*) [41], depression (*GNAI1*) [42], encephalomyopathy and brain stress response (*SLC25A42*) [43,44], and dementia/Alzheimer’s disease pathology (*SIAH3*, *SRM*, *TP53I13*) [45–47].

**Figure 5.**
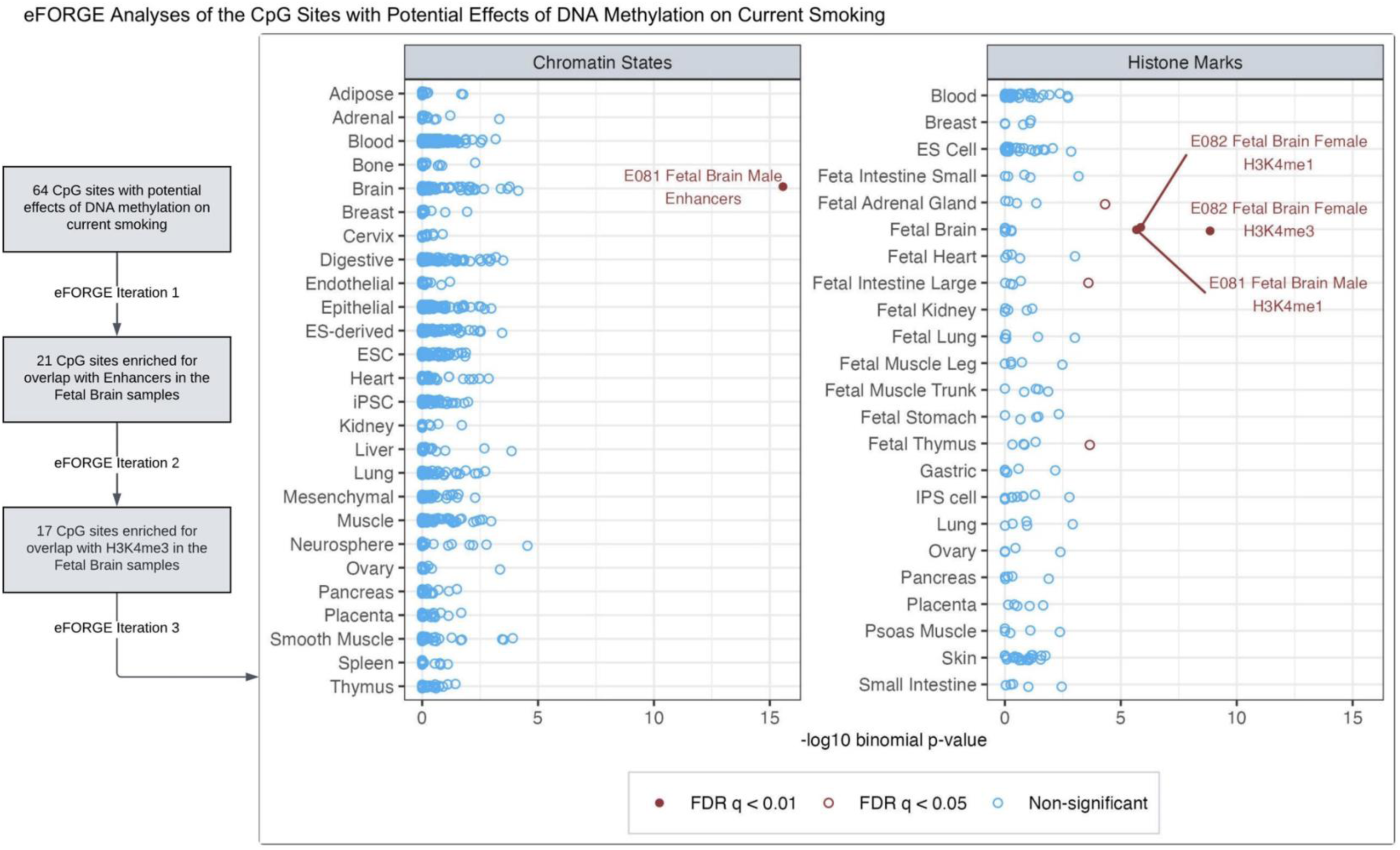
Among the CpGs with potential effects of blood DNA methylation on current smoking liability, iterative eFORGE analyses elucidated sites enriched for overlap with brain-related chromatin states and histone marks. The first iteration of eFORGE examined the 64 CpGs with potential effects of blood DNA methylation on current smoking liability (Supplementary Figure S15), revealing 21 CpGs enriched for overlap with enhancers in the brain (Supplementary Figure S18/Table S12). In follow-up analyses restricted to these 21 CpGs (eFORGE iteration 2), all 21 probes were also enriched for the brain H3K4me1 marks, while 17 of these probes overlapped with H3K4me3 marks in the brain (Supplementary Figure S22/Table S16). This iteration also showed significant enrichment (FDR q <0.01) for histone marks in other tissues, including small and large intestines, adrenal gland, and thymus. So, to identify a subset of these CpGs with potentially more specific enrichment for brain-related functional elements, we restricted further analyses to the 17 sites overlapping with the brain H3K4me3 marks (eFORGE iteration 3). This figure shows that these 17 sites showed highly specific enrichment for enhancers and histone marks in the brain (**Supplementary Tables S18-S19)**. Ten of these sites also overlapped with DNase-I hotspots in the brain (Supplementary Table S20).

Similar follow-up analyses with the CpGs overlapping with enhancers in the lung (potentially etiologically relevant tissue) and the primary B-cells in cord blood (the tissue type with the most significant enrichment) showed enrichment across several tissue/cell types, suggesting non-specificity of the overlap in these tissues (**Supplementary Figures S27-S32; Tables S21-S26**). Furthermore, the 18 CpGs overlapping with enhancers in primary B cells mapped to 16 genes, of which five have been previously associated with (any) blood cell counts, but only one with lymphocyte count in GWAS [48]. Thus, the sites driving the enrichment for B cells had little overlap with the known lymphocyte-count GWAS associations, indicating likely minimal confounding by residual cell-composition effects [35]. By comparison, the 64 CpGs with potential *DNAm → Current Smoking* effects annotated to 51 genes, of which 16 show GWAS associations with (any) blood cell counts and only two with lymphocyte count.

### Attenuated effects of former smoking on DNAm

Similar analyses for former smoking showed relatively attenuated inflation factor (λ) in all models. For instance, MR-DoC2 models fitted across the 11,124 smoking-associated CpGs had λ = 1.11 for *Former Smoking → DNAm,* and λ = 0.99 for *DNAm → Former Smoking*, compared to 1.20 and 1.01, respectively, for current smoking. Note that these λ calculations were not restricted to the former-smoking-associated CpGs to allow for a comparison with current smoking.

Among the former-smoking-associated CpGs, only five sites showed robust evidence of former smoking’s effects on DNAm, with consistent, statistically significant (FDR <0.05) causal estimates in all three models (**Supplementary Figure S33**). These CpGs include cg05575921 in *AHRR*, cg05951221, cg01940273, and cg21566642 near *ALPPL2*, and cg06126421 near *HLA-DRB5* gene (in the MHC region). The causal estimates at these sites are similar to those of *current* smoking’s effects on DNAm, with overlapping confidence intervals (**Figure 6**). Thus, the limited reversibility of smoking’s causal effects may underlie the persistent associations of former smoking with DNAm at these sites [2]. For the reverse effects of DNAm on former smoking, no CpG showed consistent (at least nominally significant) causal estimates across models (**Supplementary Figure S34**). Nevertheless, of the three CpGs with robust evidence of DNAm’s effects on current smoking, two were among the former-smoking-associated CpGs and had overlapping confidence intervals of DNAm’s estimated effects on *former* and *current* smoking (**Supplementary Figure S35**).

**Figure 6.**
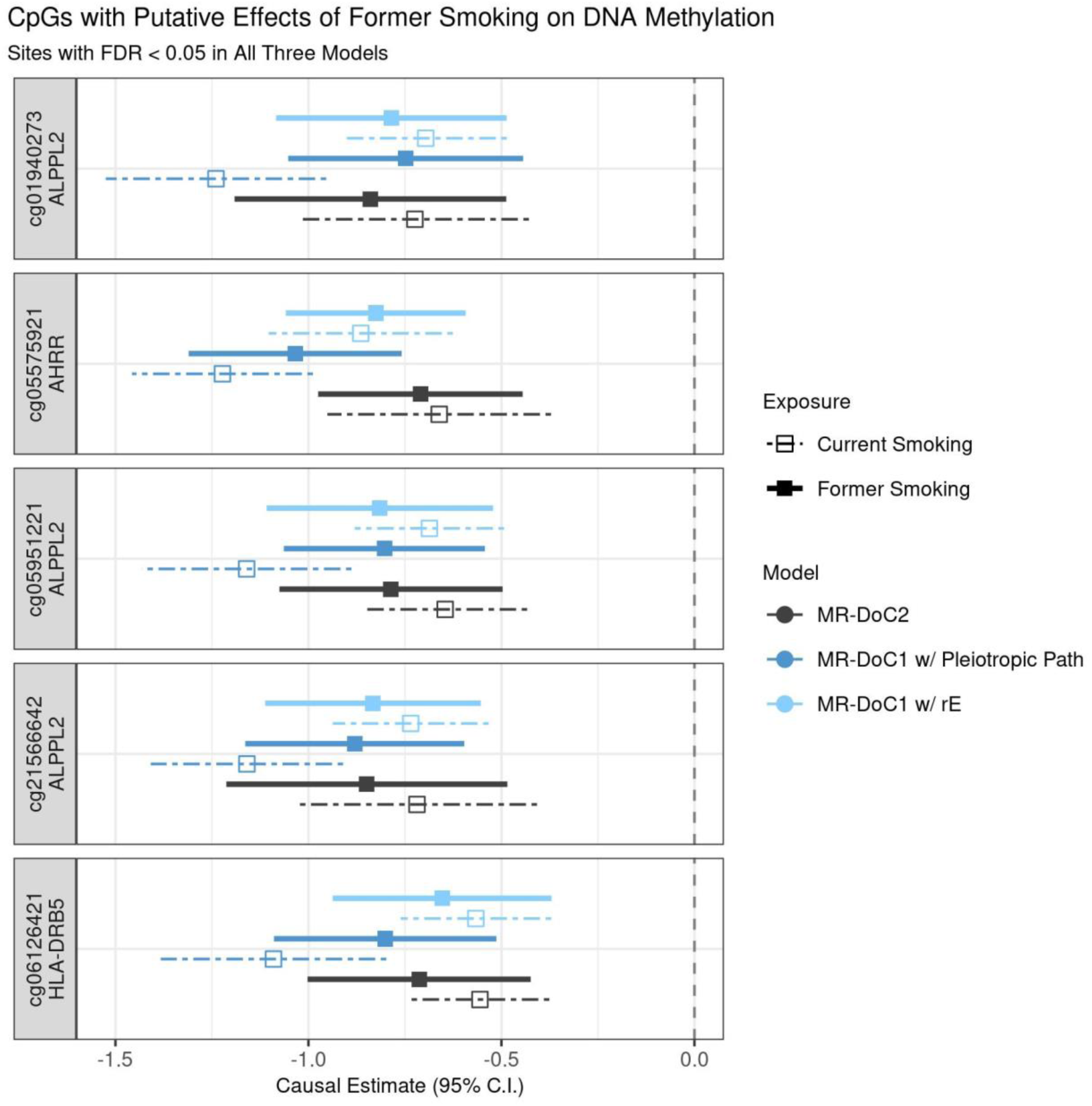
Putative causal effects of former smoking on blood DNA methylation. Estimates and Wald-type 95% confidence intervals of the causal effects of the liability for former (versus never) smoking and (residualised and standardised) DNA methylation beta-values in each of the three MR-DoC models: bidirectional MR-DoC2, MR-DoC1 with horizontal pleiotropic path, and MR-DoC1 with unique environmental confounding (rE). The corresponding estimates for current (versus never) smoking are also shown with dashed lines. The text labels on the left indicate the CpG probe IDs and the genes mapped by the CpGs. Note. The data underlying these plots are in **Supplementary Tables S1** and **S27**, indicated by the column g1_robust.

## Discussion

The integrated MR and twin models suggest that the causal effects of cigarette smoking on blood DNAm likely underlie many of the associations seen in EWAS. Compared to a handful of CpGs causally linked with smoking in previous MR analyses, we found over 500 CpGs with consistent, nominally significant effects of current smoking on DNAm. These loci show broad enrichment for tissue types and functional pathways that implicate numerous well-established harmful health outcomes of smoking, including cell- and neuro-development, carcinogenesis, and immune regulation. The discovery of more extensive and novel causal effects may partly be attributable to the study design’s ability to estimate the causal influences of *current* smoking specifically, given the considerable reversibility of most smoking-associated DNAm changes upon smoking cessation. Consistently, most of the estimated effects of smoking on DNAm were no longer significant in the analyses of former smoking. Additionally, several CpGs showed evidence of reverse and possibly bidirectional effects of DNAm on smoking liability, with a subset of these loci enriched for gene regulatory functional elements in the brain. The detection of reverse or bidirectional causal effects of blood DNAm on smoking highlights the potential utility of blood DNAm as a biomarker to monitor addiction or interventions.

Previous discordant-twin analyses in NTR found 13 CpGs with significant DNAm differences between MZ twins discordant for current smoking [24], suggesting potential causality. In the MR-DoC analyses, eight of the 13 CpGs showed robust evidence of current smoking’s effects on DNAm, while none showed reverse effects. Taken together, these findings further triangulate the evidence for smoking’s effects on DNAm at these sites. Prior summary-statistics-based MR analyses in GoDMC found no evidence of causal effects of lifetime smoking on DNAm, or *vice versa* [21]. Another study [10] applied a single MR method and found nominally significant effects of lifetime smoking on DNAm at 11 CpGs from the Illumina MethylationEPIC array [49], of which two (cg14580211, cg15212295) overlap with Illumina 450k array data used in the current study. In our MR-DoC analyses, only cg14580211 showed replication in the form of consistent negative causal estimates of current smoking on DNAm. Furthermore, the nine CpGs with previously reported reverse effects of DNAm on lifetime smoking behaviour in a single MR model [11] showed inconsistent estimates in the three MR-DoC models. Interestingly, two of these CpGs (cg09099830 and cg24033122; both in gene *ITGAL*) showed consistent, nominally significant effects of current smoking on DNAm, underscoring the need for further replication of both prior and current findings.

Of the three loci with robust evidence of DNAm’s effects on current smoking liability, two are located in genes *GNG7* and *RGS3,* which are integral to G protein-coupled receptor (GPCR) signalling, adding to the growing literature on GPCR signalling pathways’ potential role in behavioural and neuropsychiatric outcomes [50]. Specifically, differential expression of both *GNG7* [51] and *RGS3* [52] has been associated with addiction-related phenotypes in model organisms. The third CpG annotates to *SLC15A4*, which encodes a lysosomal peptide/histidine transporter involved in antigen presentation and innate immune response [53], including in mast cells [54]. Thus, DNAm variation at this locus may reflect individual differences in immunological tolerance of cigarette smoke and, consequently, maintenance of smoking behaviour. Interestingly, these CpGs were significantly associated with neither cannabis use [7] nor alcohol consumption [6] in recent large-scale EWASs. However, these studies reported DNAm associations conditional on cigarette smoking, making them unsuitable for gauging whether the CpGs with putative effects on smoking liability are also associated with other substances. This raises the question of whether cigarette smoking should always be used as a covariate in EWAS. If so, it may be prudent to report supplementary EWAS results without smoking as a covariate, as some CpGs may have reverse or bidirectional causal relationships with smoking.

Several factors need to be considered when interpreting the above results. We analysed DNAm from whole blood, but smoking’s causal relationships with DNAm may differ between specific blood cell types. The results may also vary in other peripheral tissues, like buccal cells [55], and other tissues relevant to smoking, like the brain. Moreover, the highly variable predictive strength of mQTL allelic scores across CpGs (incremental-R^2^ range: 0.43-76.95%; median 4.61%) affected the power to detect causal effects of blood DNAm on smoking liability [38].

When considering similar model applications across different health traits, this impact on power is relevant to both directions of causation, as the IV of other traits may not be as strong as the smoking PRS. Additionally, the Illumina 450k microarray used in this study covers a small fraction of genome-wide potential methylation sites. Moreover, many of the measured smoking-associated CpGs lacked a “relevant” mQTL allelic score with F-statistic >10 (**Supplementary Figure S36**), and so have yet to be tested for *DNAm → Smoking* causal effects. Newer low-cost sequencing technology [56] may facilitate further causal discovery in the future.

Like all MR studies, the current results depend on the validity of the IV assumptions [28], which can be difficult to test. Here, we relied on the statistical significance and consistency of the causal estimates across different MR-DoC model specifications to account for potential assumption violations, particularly horizontal pleiotropy. Yet, we cannot rule out residual bias due to violations of the assumptions underlying MR [28] and twin modelling [57]. Moreover, current MR-DoC models estimated linear causal effects. However, since DNAm is constrained within certain biologically plausible values, the impact of smoking on DNAm may depend on *prior* DNAm. Further development of MR-DoC models with interaction or quadratic effects will benefit the study of such non-linear causal effects. Finally, we examined causality using only binary smoking-status variables, as the number of individuals endorsing current or former smoking was too small to fit MR-DoC models to smoking quantity (e.g., cigarettes per day) or time since quitting. Further research with larger samples is needed to examine such dose-response causal relationships.

The current study included participants of European ancestry only. Although prior EWASs show highly concordant associations across ancestries [2,58], examining the generalizability of causal estimates in non-European populations is an essential objective for further research. As MR-DoC models estimate causal effects specific to the target dataset, rather than the discovery GWAS samples, future research may apply this study design to subpopulations of interest, e.g., stratified by sex or age, provided the population-wide GWAS results generalise adequately. Future applications of MR-DoC analyses to DNAm data may also extend the current work to other traits that show robust associations with DNAm [59] and have strong genetic IVs.

In conclusion, the inability to establish causality is a clear limitation of EWAS based on surrogate tissues such as blood. Here, we applied the MR-DoC designs to examine the causality between cigarette smoking and blood DNAm. The results suggest that many of the EWAS associations are likely driven by the causal effects of current smoking on DNAm, and provide evidence for reverse and potentially bidirectional causal relationships at some sites. Underscoring the continuing value of twin studies for health and behaviour [60], our study highlights the value of integrating DNAm, phenotypic information, and genetic data in twin studies to uncover causal relationships of peripheral blood DNAm with complex traits. This study design might be valuable for detecting causal epigenetic biomarkers of mental health in general.

## Acknowledgements

NTR warmly thanks all participants. Epigenetic data were generated at the Human Genomics Facility (HuGe-F) at ErasmusMC Rotterdam (http://www.glimdna.org/) as part of the Biobank-based Integrative Omics Study Consortium. We thank Dr. Scott Vrieze (University of Minnesota) for providing the leave-one-out GWAS summary statistics from the GWAS & Sequencing Consortium of Alcohol and Nicotine Use (GSCAN).

## Data Availability

Data from the Netherlands Twin Register (NTR) may be accessed for research purposes by submitting a data-sharing request. Further information about NTR data access is available at https://ntr-data-request.psy.vu.nl/.

Results of all MR-DoC models fitted in this study are available as Supplementary Data on OSF (doi:10.17605/OSF.IO/R6HVY).

## Code Availability

The code used in the analyses for this study is available at: https://github.com/singh-madhur/MRDOC_Smoking_DNAm_NTR.

## References

1. Wei S, Tao J, Xu J, Chen X, Wang Z, Zhang N, et al. Ten Years of EWAS. Adv Sci. 2021 Oct 1;8(20):2100727.

2. Joehanes R, Just AC, Marioni R, Pilling L, Reynolds L, Mandaviya PR, et al. Epigenetic Signatures of Cigarette Smoking. Circ Cardiovasc Genet. 2016;9(5):436–47.

3. Lawlor DA, Harbord RM, Sterne JA, Timpson N, Davey Smith G. Mendelian randomization: using genes as instruments for making causal inferences in epidemiology. Stat Med. 2008 Apr 15;27(8):1133–63.

4. Zillich L, Poisel E, Streit F, Frank J, Fries GR, Foo JC, et al. Epigenetic Signatures of Smoking in Five Brain Regions. J Pers Med. 2022;12(4):566.

5. Hannon E, Dempster E, Viana J, Burrage J, Smith AR, Macdonald R, et al. An integrated genetic-epigenetic analysis of schizophrenia: evidence for co-localization of genetic associations and differential DNA methylation. Genome Biol. 2016 Aug 30;17(1):176.

6. Dugué PA, Wilson R, Lehne B, Jayasekara H, Wang X, Jung CH, et al. Alcohol consumption is associated with widespread changes in blood DNA methylation: Analysis of cross-sectional and longitudinal data. Addict Biol. 2021 Jan 1;26(1):e12855.

7. Nannini DR, Zheng Y, Joyce BT, Kim K, Gao T, Wang J, et al. Genome-wide DNA methylation association study of recent and cumulative marijuana use in middle aged adults. Mol Psychiatry [Internet]. 2023 May 31; Available from: 10.1038/s41380-023-02106-y

8. Dhana K, Braun KVE, Nano J, Voortman T, Demerath EW, Guan W, et al. An Epigenome-Wide Association Study of Obesity-Related Traits. Am J Epidemiol. 2018 Aug 1;187(8):1662–9.

9. Davey Smith G, Ebrahim S. Mendelian randomization: Can genetic epidemiology contribute to understanding environmental determinants of disease? Int J Epidemiol. 2003 Feb;32(1):1–22.

10. Sun YQ, Richmond RC, Suderman M, Min JL, Battram T, Flatberg A, et al. Assessing the role of genome-wide DNA methylation between smoking and risk of lung cancer using repeated measurements: the HUNT study. Int J Epidemiol. 2021;50(5):1482–97.

11. Jamieson E, Korologou-Linden R, Wootton RE, Guyatt AL, Battram T, Burrows K, et al. Smoking, DNA Methylation, and Lung Function: A Mendelian Randomization Analysis to Investigate Causal Pathways. Am J Hum Genet. 2020 Mar 5;106(3):315–26.

12. Burgess S, Thompson SG, CRP CHD Genetics Collaboration. Avoiding bias from weak instruments in Mendelian randomization studies. Int J Epidemiol. 2011 Jun;40(3):755–64.

13. Dugué PA, Jung CH, Joo JE, Wang X, Wong EM, Makalic E, et al. Smoking and blood DNA methylation: an epigenome-wide association study and assessment of reversibility. Epigenetics. 2020 Apr 2;15(4):358–68.

14. Heath AC, Kessler RC, Neale MC, Hewitt JK, Eaves LJ, Kendler KS. Testing hypotheses about direction of causation using cross-sectional family data. Behav Genet. 1993 Jan 1;23(1):29–50.

15. Minică CC, Dolan CV, Boomsma DI, De Geus E, Neale MC. Extending Causality Tests with Genetic Instruments: An Integration of Mendelian Randomization with the Classical Twin Design. Behav Genet. 2018;48(4):337–49.

16. Castro-de-Araujo LFS, Singh M, Zhou Y, Vinh P, Verhulst B, Dolan CV, et al. MR-DoC2: Bidirectional Causal Modeling with Instrumental Variables and Data from Relatives. Behav Genet. 2023 Feb 1;53(1):63–73.

17. Minică CC, Boomsma DI, Dolan CV, De Geus E, Neale MC. Empirical comparisons of multiple Mendelian randomization approaches in the presence of assortative mating. Int J Epidemiol. 2020 Aug 1;49(4):1185–93.

18. Ligthart L, van Beijsterveldt CEM, Kevenaar ST, de Zeeuw E, van Bergen E, Bruins S, et al. The Netherlands Twin Register: Longitudinal Research Based on Twin and Twin-Family Designs. Twin Res Hum Genet. 2019;22(6):623–36.

19. Willemsen G, de Geus EJC, Bartels M, van Beijsterveldt CEMT, Brooks AI, Estourgie-van Burk GF, et al. The Netherlands Twin Register Biobank: A Resource for Genetic Epidemiological Studies. Twin Res Hum Genet. 2012/02/21 ed. 2010;13(3):231–45.

20. Singh M, Verhulst B, Vinh P, Zhou Y (Daniel), Castro-de-Araujo LFS, Hottenga JJ, et al. Using Instrumental Variables to Measure Causation over Time in Cross-Lagged Panel Models. Multivar Behav Res. 2024 Feb 15;59(2):342–70.

21. Min JL, Hemani G, Hannon E, Dekkers KF, Castillo-Fernandez J, Luijk R, et al. Genomic and phenotypic insights from an atlas of genetic effects on DNA methylation. Nat Genet. 2021 Sep 1;53(9):1311–21.

22. Bibikova M, Barnes B, Tsan C, Ho V, Klotzle B, Le JM, et al. High density DNA methylation array with single CpG site resolution. New Genomic Technol Appl. 2011 Oct 1;98(4):288–95.

23. van Dongen J, Nivard MG, Willemsen G, Hottenga JJ, Helmer Q, Dolan CV, et al. Genetic and environmental influences interact with age and sex in shaping the human methylome. Nat Commun. 2016 Sep 1;7(1):11115.

24. van Dongen J, Willemsen G, BIOS Consortium, de Geus EJ, Boomsma DI, Neale MC. Effects of smoking on genome-wide DNA methylation profiles: A study of discordant and concordant monozygotic twin pairs. Aldrich M, Rathmell WK, Aldrich M, Craig J, Kaprio J, editors. eLife. 2023 Aug 10;12:e83286.

25. Chang CC, Chow CC, Tellier LC, Vattikuti S, Purcell SM, Lee JJ. Second-generation PLINK: rising to the challenge of larger and richer datasets. GigaScience. 2015 Dec 1;4(1).

26. Vilhjálmsson J, Yang J, Finucane K, Gusev A, Lindström S, Ripke S, et al. Modeling Linkage Disequilibrium Increases Accuracy of Polygenic Risk Scores. Am J Hum Genet. 2015 Oct 1;97(4):576–92.

27. Saunders GRB, Wang X, Chen F, Jang SK, Liu M, Wang C, et al. Genetic diversity fuels gene discovery for tobacco and alcohol use. Nature. 2022 Dec 22;612(7941):720–4.

28. Richmond RC, Davey Smith G. Mendelian Randomization: Concepts and Scope. Cold Spring Harb Perspect Med. 2022 Jan 1;12(1):a040501.

29. Neale MC, Hunter MD, Pritikin JN, Zahery M, Brick TR, Kirkpatrick RM, et al. OpenMx 2.0: Extended Structural Equation and Statistical Modeling. Psychometrika. 2016;81(2):535–49.

30. Verhulst B, Neale MC. Best Practices for Binary and Ordinal Data Analyses. Behav Genet. 2021;51(3):204–14.

31. van Iterson M, van Zwet EW, Heijmans BT, the BIOS Consortium. Controlling bias and inflation in epigenome- and transcriptome-wide association studies using the empirical null distribution. Genome Biol. 2017 Jan 27;18(1):19.

32. Gogarten SM, Bhangale T, Conomos MP, Laurie CA, McHugh CP, Painter I, et al. GWASTools: an R/Bioconductor package for quality control and analysis of genome-wide association studies. Bioinformatics. 2012 Dec 1;28(24):3329–31.

33. Benjamini Y, Hochberg Y. Controlling the False Discovery Rate: A Practical and Powerful Approach to Multiple Testing. J R Stat Soc Ser B Methodol. 1995 Jan 1;57(1):289–300.

34. Zhou Y, Zhou B, Pache L, Chang M, Khodabakhshi AH, Tanaseichuk O, et al. Metascape provides a biologist-oriented resource for the analysis of systems-level datasets. Nat Commun. 2019 Apr 3;10(1).

35. Breeze CE, Paul DS, van Dongen J, Butcher LM, Ambrose JC, Barrett JE, et al. eFORGE: A Tool for Identifying Cell Type-Specific Signal in Epigenomic Data. Cell Rep. 2016 Nov 15;17(8):2137–50.

36. Breeze CE, Reynolds AP, van Dongen J, Dunham I, Lazar J, Neph S, et al. eFORGE v2.0: updated analysis of cell type-specific signal in epigenomic data. Bioinformatics. 2019 Nov 15;35(22):4767–9.

37. Breeze CE. Cell Type-Specific Signal Analysis in Epigenome-Wide Association Studies. In: Guan W, editor. Epigenome-Wide Association Studies: Methods and Protocols [Internet]. New York, NY: Springer US; 2022. p. 57–71. Available from: 10.1007/978-1-0716-1994-0_5

38. Castro-de-Araujo LF, Singh M, Zhou Y, Vinh P, Maes HH, Verhulst B, et al. Power, measurement error, and pleiotropy robustness in twin-design extensions to Mendelian Randomization. Research square. United States; 2023. p. rs.3.rs-3411642.

39. Edenberg HJ, Koller DL, Xuei X, Wetherill L, McClintick JN, Almasy L, et al. Genome-Wide Association Study of Alcohol Dependence Implicates a Region on Chromosome 11. Alcohol Clin Exp Res. 2010 May 1;34(5):840–52.

40. Khan AH, Bagley JR, LaPierre N, Gonzalez-Figueroa C, Spencer TC, Choudhury M, et al. Genetic pathways regulating the longitudinal acquisition of cocaine self-administration in a panel of inbred and recombinant inbred mice. Cell Rep. 2023 Aug 29;42(8):112856.

41. Jin X, Dong S, Yang Y, Bao G, Ma H. Nominating novel proteins for anxiety via integrating human brain proteomes and genome-wide association study. J Affect Disord. 2024 Aug 1;358:129–37.

42. Sarkar A, Chachra P, Kennedy P, Pena CJ, Desouza LA, Nestler EJ, et al. Hippocampal HDAC4 Contributes to Postnatal Fluoxetine-Evoked Depression-Like Behavior. Neuropsychopharmacology. 2014 Aug 1;39(9):2221–32.

43. Aldosary M, Baselm S, Abdulrahim M, Almass R, Alsagob M, AlMasseri Z, et al. SLC25A42-associated mitochondrial encephalomyopathy: Report of additional founder cases and functional characterization of a novel deletion. JIMD Rep. 2021 Jul 1;60(1):75– 87.

44. Stankiewicz AM, Jaszczyk A, Goscik J, Juszczak GR. Stress and the brain transcriptome: Identifying commonalities and clusters in standardized data from published experiments. Prog Neuropsychopharmacol Biol Psychiatry. 2022 Dec 20;119:110558.

45. Cochran JN, Acosta-Uribe J, Esposito BT, Madrigal L, Aguillón D, Giraldo MM, et al. Genetic associations with age at dementia onset in the PSEN1 E280A Colombian kindred. Alzheimers Dement. 2023 Sep 1;19(9):3835–47.

46. Mahajan UV, Varma VR, Griswold ME, Blackshear CT, An Y, Oommen AM, et al. Dysregulation of multiple metabolic networks related to brain transmethylation and polyamine pathways in Alzheimer disease: A targeted metabolomic and transcriptomic study. PLOS Med. 2020 Jan 24;17(1):e1003012.

47. Blanco-Luquin I, Acha B, Urdánoz-Casado A, Sánchez-Ruiz De Gordoa J, Vicuña-Urriza J, Roldán M, et al. Early epigenetic changes of Alzheimer’s disease in the human hippocampus. Epigenetics. 2020 Oct 2;15(10):1083–92.

48. Vuckovic D, Bao EL, Akbari P, Lareau CA, Mousas A, Jiang T, et al. The Polygenic and Monogenic Basis of Blood Traits and Diseases. Cell. 2020 Sep 3;182(5):1214–1231.e11.

49. Pidsley R, Zotenko E, Peters TJ, Lawrence MG, Risbridger GP, Molloy P, et al. Critical evaluation of the Illumina MethylationEPIC BeadChip microarray for whole-genome DNA methylation profiling. Genome Biol. 2016 Oct 7;17(1):208.

50. Wong TS, Li G, Li S, Gao W, Chen G, Gan S, et al. G protein-coupled receptors in neurodegenerative diseases and psychiatric disorders. Signal Transduct Target Ther. 2023 May 3;8(1):177.

51. Stankiewicz AM, Goscik J, Dyr W, Juszczak GR, Ryglewicz D, Swiergiel AH, et al. Novel candidate genes for alcoholism — transcriptomic analysis of prefrontal medial cortex, hippocampus and nucleus accumbens of Warsaw alcohol-preferring and non-preferring rats. Pharmacol Biochem Behav. 2015 Dec 1;139:27–38.

52. Burchett SA, Bannon MJ, Granneman JG. RGS mRNA Expression in Rat Striatum. J Neurochem. 1999 Apr 1;72(4):1529–33.

53. Chen X, Xie M, Zhang S, Monguió-Tortajada M, Yin J, Liu C, et al. Structural basis for recruitment of TASL by SLC15A4 in human endolysosomal TLR signaling. Nat Commun. 2023 Oct 20;14(1):6627.

54. Kobayashi T, Tsutsui H, Shimabukuro-Demoto S, Yoshida-Sugitani R, Karyu H, Furuyama-Tanaka K, et al. Lysosome biogenesis regulated by the amino-acid transporter SLC15A4 is critical for functional integrity of mast cells. Int Immunol. 2017 Dec 31;29(12):551–66.

55. Teschendorff AE, Yang Z, Wong A, Pipinikas CP, Jiao Y, Jones A, et al. Correlation of Smoking-Associated DNA Methylation Changes in Buccal Cells With DNA Methylation Changes in Epithelial Cancer. JAMA Oncol. 2015 Jul 1;1(4):476–85.

56. Simpson JT, Workman RE, Zuzarte PC, David M, Dursi LJ, Timp W. Detecting DNA cytosine methylation using nanopore sequencing. Nat Methods. 2017 Apr 1;14(4):407–10.

57. Evans DM, Gillespie NA, Martin NG. Biometrical genetics. Biol Psychol. 2002;61(1):33– 51.

58. Fang F, Quach B, Lawrence KG, van Dongen J, Marks JA, Lundgren S, et al. Trans-ancestry epigenome-wide association meta-analysis of DNA methylation with lifetime cannabis use. Mol Psychiatry [Internet]. 2023 Nov 7; Available from: 10.1038/s41380-023-02310-w

59. Jin Z, Liu Y. DNA methylation in human diseases. Genes Dis. 2018 Mar 1;5(1):1–8.

60. Hagenbeek FA, Hirzinger JS, Breunig S, Bruins S, Kuznetsov DV, Schut K, et al. Maximizing the value of twin studies in health and behaviour. Nat Hum Behav. 2023 Jun 1;7(6):849–60.

