## Supplementary Information for "Unidirectional and Bidirectional Causation between Smoking and Blood DNA Methylation: Evidence from Twin-based Mendelian Randomisation"

### Supplementary Methods

In this study, we analyzed data from the Netherlands Twin Register (NTR) [1] to examine the causal influences between smoking status and blood DNA methylation (DNAm) using MR-DoC models [2,3]. In the current analyses, we included data from European-ancestry adult twins with both genotypic and DNAm data, comprising 2,577 individuals (67% female).

#### Genotypic Data, Principal Components Analysis, and Ancestry Outlier Pruning

The DNA samples included in the current study were genotyped on 3 SNP (single nucleotide polymorphism) microarray platforms: Affymetrix 6.0 (N= 2,399), Affymetrix Axiom (N= 83), and Illumina GSA NTR array (N= 95). Genotype calling was done following the manufacturer's protocols. Sample and variant quality control (QC), imputation, genetic principal component analysis (PCA), and ancestry assignment have been previously described [4]. Briefly, after QC and harmonizing variants across the three platforms, the data were aligned to the positive strand of *Genome Reference Consortium Human Build 37* (GRCh37) and then imputed against the European (EUR) super-population of the 1000 Genomes Project Phase-3 (KGP3) [5], the Haplotype Reference Consortium (HRC) [6] 1.1 (Ega version), and the Genome of the Netherlands Consortium (GoNL) [7] reference panels. Using SmartPCA in EIGENSTRAT [8], the first 20 PCs for the genotypic data were calculated in the KGP3 data, and the NTR samples were then projected onto the PC space based on the SNP weights. Samples identified as outliers in the PC space were then excluded.

#### Peripheral Blood DNA Methylation Data

Epigenome-wide DNAm in peripheral whole blood was measured with the Infinium HumanMethylation450 BeadChip Kit (i.e., the Illumina 450k microarray), following the manufacturer's protocol [9]. QC and normalization of the DNAm data were performed using a custom pipeline developed by the BIOS (Biobank-based Integrative Omics Study) Consortium, as previously described [10]. Briefly, sample QC was done using MethylAid [11], followed by probe QC with DNAmArray [12]. The latter removed the probes with a raw signal intensity of zero, bead number <3, or a detection p-value >0.01, as well as the ambiguously mapped probes. Next, samples and probes with >5% missingness were removed. The resulting DNAm data were normalized using the Functional normalization algorithm [13] implemented in DNAmArray [12], with the first four PCs (with eigenvalue >1) derived from control probes. Finally, the probes containing a SNP within the CpG site (at C or G nucleotide) were removed regardless of the minor allele frequency. These SNPs were previously identified using DNA sequencing data from the Dutch population in GoNL [7]. For the current analyses, only autosomal probes were included, yielding 411,169 CpG sites that passed all QC metrics.

#### Smoking Assessment at Blood Sampling

Self-reported cigarette smoking status was recorded through an interview during the home visit for blood sample collection in 2004-2008 and 2010-2011. Participants were asked, “Do you smoke?” with one of three possible answers: “No, I never smoked” (N = 1,492), “No, but I did in the past” (N = 549), and “Yes” (N = 528). Those endorsing current smoking were asked how many years they had been smoking and how many cigarettes or rolling tobacco they smoked per day. Those endorsing former smoking were asked how many years ago they quit smoking, how many years they had smoked before quitting, and the maximum number of cigarettes or rolling tobacco they used to smoke per day. The original wording in Dutch is shown below.

| | | |
|---|---|---|
| Rookt u? | 1. Ja | 1a: hoelang rookt u al? ……………………..jaar |
| | | 1b: hoeveel sigaretten/ shagjes per dag?<br>……………………………..sigaretten/ shagjes |
| | 2. Nee, wel in het verleden | 2a: hoelang is dat geleden?.........................jaar |
| | | 2b: hoeveel jaren heeft u gerookt?..............jaar |
| | | 2c: hoeveel rookte u per dag (max)?<br>...……………………..……..sigaretten/ shagjes |
| | 3. Nooit | |
| Gerookt binnen laatste uur voor bloedafname? | 1. Ja<br>2. Nee<br>3. Nvt | |

The responses were checked for consistency with the information from the NTR longitudinal surveys filled out closest to blood sampling. As previously described [14], potential misclassification of smoking status through self-reports was evaluated based on plasma cotinine levels (a metabolite of nicotine and a biomarker of smoking exposure), measured in a subset of the sample. Of the 591 individuals with self-reported never smoking and measured plasma cotinine, only five (0.8%) had cotinine levels indicative of smoking (≥15 ng/ml), thus suggesting low misclassification of smoking status. The number of individuals endorsing current or former smoking was too small to evaluate a dose-response relationship of the causal effects in MR-DoC models restricted to currently or formerly smoking individuals. Likewise, the sample with former smoking was too small to examine the effect of “time since quitting smoking” on DNAm.

#### mQTL Allelic Score

We identified 12,940 smoking-associated CpGs with *cis*-mQTL summary statistics available from GoDMC [15] (excluding NTR), using GoDMC’s definition of “*cis*” interval (within 1Mb of the CpG). In GoDMC, the contributing cohorts performed genome-wide mQTL analyses, testing the associations of ~480,000 CpG sites with ~12 million SNPs. However, before the meta-analysis, the cohort-level results were filtered to retain the SNP-CpG pairs with $p < 1 \times 10^{-5}$

within the cohort. Thus, since the summary statistics were already partly thresholded, we computed the mQTL allelic scores by applying clumping and thresholding in *PLINK1.9* [16], using summary statistics from the Genetics of DNA Methylation Consortium (GoDMC; excluding NTR) [15]. Linkage disequilibrium (LD)-based clumping was performed using `--clump-p1 1 --clump-kb 250`, with two levels of LD $r^2$ (0.5 and 0.1) specified for `--clump-r2`, thus yielding two sets of LD-clumped *cis*-SNPs. Using either set of SNPs, we computed the allelic score with `--score` at a threshold of 0.05 (applied with `--q-score-range`). If none of the SNPs had $p < 0.05$, no threshold was applied for score calculation. An additional allelic score was calculated using the top *cis*-mQTL (with the minimum association p-value) for each CpG. Thus, for every CpG, three scores were calculated (two LD-clumped mQTL allelic scores, plus the top-mQTL), though these scores were not necessarily distinct; for example, if a CpG had only one *cis*-SNP, all three criteria yielded the same score. Likewise, for some CpGs, the two LD-clumping cut-offs resulted in the same set of SNPs and, hence, identical mQTL allelic scores.

To assess the strength of an mQTL allelic score, we first estimated its incremental $R^2$ by fitting generalized estimating equations (GEE), controlling for the standard EWAS covariates (as above), genotyping platform, and the first ten genetic PCs. For each CpG, the mQTL allelic score with the highest incremental $R^2$ was retained for further filtering based on F-statistic. For each CpG, the effective GEE sample size ($N_{Eff}$) was computed using the following formulae:

$$N_{Eff}^{MZ} = \frac{2 * N_{MZ}}{1 + r_{MZ}}$$

$$N_{Eff}^{DZ} = \frac{2 * N_{DZ}}{1 + r_{DZ}}$$

$$N_{Eff} = N_{Eff}^{MZ} + N_{Eff}^{DZ} + N_{Ind}$$

where, $N_{Eff}^{MZ}$ and $N_{Eff}^{DZ}$ are the estimated effective sample sizes of MZ and DZ twins, $N_{MZ}$ and $N_{DZ}$ are the numbers of complete MZ and DZ twin pairs, while $r_{MZ}$ and $r_{DZ}$ are the twin phenotypic (DNAm) correlations in MZ and DZ twin pairs, respectively. $N_{Ind}$ is the number of individuals without the co-twin.
The estimated effective sample size was then used to transform the incremental $R^2$ value into an F-statistic as:

$$F = \frac{R^2}{1 - R^2} \times \frac{N_{Eff} - K}{K - 1}$$

where $K = 2$, given two parameter estimates: the intercept and the regression coefficient of the mQTL allelic score.

#### Polygenic Risk Score of Smoking

The PRS of smoking was based on the European-ancestry summary statistics from the genome-wide association study (GWAS) of smoking initiation (lifetime regular smoking) by GSCAN (GWAS & Sequencing Consortium of Alcohol and Nicotine use)[17], excluding the NTR from the meta-analysis.

As described in a previous study using the same PRS in the NTR[4], the post-imputation SNPs from the merged best-guess three-platform data were QCed to satisfy the following criteria: MAF >0.01, HWE p >0.00001, Mendel error rate < 1%, and genotype call rate over 98%. Furthermore, the imputation info for the three platforms needed to be above 0.10, and the allele frequency between platforms after imputation could not differ more than 2%, leaving a total of 7,551,860 post-QC SNPs for analysis. The PRS was calculated using *LDpred* v0.9[18], with HRC+GoNL as the LD (linkage disequilibrium) reference panel. For estimating the target LD structure, we used a subset of unrelated individuals and a set of well-imputed variants in the NTR. The parameter `ld_radius` was set by dividing the number of variants in common (from the output of the coordination step) by 12000. For the coordination step, the median sample size was used as the input value for N. For the LDpred step, we applied the following thresholds for the fraction of variants with non-zero effects (in addition to the default infinitesimal model): `--PS=0.5,0.3,0.2,0.1,0.05,0.01`.

To determine the LDPred threshold that yielded the PRS with the highest predictive power for the variables of interest (current vs. never and former vs. never smoking), we fitted logistic regression models in R (v4.3.2) to estimate incremental $R^2$ on a liability scale. We first fitted a null logistic regression model using the `glm()` function with `family=binomial(link='logit')` and a standard set of covariates comprising age (linear and quadratic), sex, SNP microarray platform (dummy variables), and the first ten genetic PCs (without including the PRS). Then, we fitted a full model with the PRS as an additional independent variable. We estimated the liability-scale $R^2$ in both models and then the difference in the two $R^2$ estimates as the variance in the outcome variable explained by the PRS (controlling for the covariates). For both outcome variables (current and former smoking), the PRS with the highest incremental $R^2$ was based on a threshold of 0.1 and thus retained for further analyses. The PRS was residualized for the SNP microarray platform and the first ten genetic PCs using linear regression models. The residuals were then standardized to have a mean of zero and an S.D. of one before using it as an IV in the MR-DoC models.

#### Univariate Twin Models

Before fitting the MR-DoC models, we examined univariate ACE twin models of smoking status to estimate the additive genetic (A), shared environmental (C), and unique environmental (E) variance components of the latent liability scale, with age and sex as covariates. Maximum-likelihood tetrachoric correlation estimates for current versus never smoking were: $r_{MZ} = 0.925$ ($S.E. = 0.021$) in MZ pairs, and $r_{DZ} = 0.533$ ($S.E. = 0.083$) in DZ pairs. Likewise, former versus never smoking had $r_{MZ} = 0.822$ ($S.E. = 0.038$) and $r_{DZ} = 0.474$ ($S.E. = 0.096$). Based on likelihood-ratio tests (LRT), an AE twin model was the most parsimonious model for both current versus never (AE versus ACE LRT $p = 0.417$) and former versus never smoking (AE versus ACE LRT $p = 0.530$) (**Supplementary Table S31**). The estimated variance components of current versus never smoking liability were $A = 0.927$ (maximum-likelihood 95% confidence interval: 0.879, 0.959) and $E = 0.073$ (0.041, 0.121). The corresponding estimates of former versus never smoking were $A = 0.827$ (0.745, 0.888) and $E = 0.173$ (0.112, 0.255).

Prior twin analyses of DNAm at CpG sites in NTR [10] showed that, of the 411,169 autosomal post-QC CpG sites, the AE twin model was the best fitting model at all but 426 sites, with significant (after multiple-testing correction of LRT p-values) C variance at 185 sites and significant non-additive genetic (D) variance at 241 sites. Of the smoking-associated CpGs [19], only two CpGs had significant estimates of C, while only seven CpGs had significant estimates of D. Thus, in the MR-DoC models, we specified an AE variance decomposition of DNAm at all smoking-associated CpGs. Note that, in the results presented in the main text, none of the CpG sites with consistent, nominally significant estimates of causal effects in either direction (525 sites with *current smoking → DNAm*; 64 sites with *DNAm → current smoking*) have significant C or D estimates per the previous univariate twin analyses [10]. Moreover, since smoking status liability also has an AE variance decomposition, including a C or D variance component of DNAm in the model would not change the possible sources of covariance between smoking status and DNAm in the model.

#### MR-DoC Models

We used the *OpenMx* (version 2.21.8) [20] package in R (version 4.3.2) to fit the MR-DoC models using the code provided in the original publications [2,3]. Binary smoking status was examined under the liability threshold model [21], assuming a latent liability distribution with its mean fixed at zero and variance fixed at one, while the threshold was freely estimated.

In each MR-DoC model, the residual variance of smoking status liability is decomposed into $a_S^2$ (A) and $e_S^2$ (E), while that of DNAm is decomposed into $a_D^2$ (A) and $e_D^2$ (E). The correlation between the latent A factors of smoking and DNAm (rA) represents the confounding due to additive genetic factors. The correlation between the latent E factors (rE) represents the confounding due to unique environmental factors. Across all models, the causal path from smoking to DNAm is labeled $g_1$, while that from DNAm to smoking is labeled $g_2$. The residualized PRS and mQTL allelic scores are regressed on respective latent factors, representing the underlying "true" standardized scores with mean fixed at zero and variance fixed at one. The coefficient of the path from the latent score to the observed score estimates the standard deviation of the observed score ($SD_{PRS}$ and $SD_{mQTL}$, respectively).

We fitted five sets of MR-DoC models with current versus never smoking and similar sets with former versus never smoking (**Figure 1**): (1) *Smoking → DNAm* MR-DoC1 with horizontal pleiotropy, (2) *Smoking* → *DNAm* MR-DoC1 with unique environmental confounding, (3) *DNAm* → *Smoking* MR-DoC1 with horizontal pleiotropy, (4) *DNAm* → *Smoking* MR-DoC1 with unique environmental confounding, and (5) bidirectional MR-DoC2. Each model included age and sex as covariates of smoking status. Thus, for each CpG site included in the analyses, three causal estimates were obtained in either direction (*Smoking* → *DNAm*, or *DNAm* → *Smoking*) from (1) MR-DoC1 with horizontal pleiotropy, (2) MR-DoC1 with unique environmental confounding, and (3) MR-DoC2. For each set of causal estimates across CpG sites, we calculated the Bayesian inflation factor (λ) using the R package *bacon* [22], made QQ plots using the R package *GWASTools* [23], and then applied Benjamini-Hochberg FDR correction [24] to the p-values using the R package *qvalue* [25]. For Bonferroni multiple-testing correction, the significance level was defined as $\alpha = 0.05/16940 = 2.95 \times 10^{-6}$ for *Current Smoking* → *DNAm* MR-DoC1 models and $\alpha = 0.05/11124 = 4.49 \times 10^{-6}$ for *DNAm* → *Current Smoking* MR-DoC1 and bidirectional current-smoking MR-DoC2 models.

#### Functional Enrichment Analyses

We used Metascape [26] (v3.5.20240101; https://metascape.org/gp/index.html#/main/step1, with the default settings for "Express" analyses) to perform gene-set annotation and functional enrichment analyses of the CpGs with potential causal effects in either direction. The input list of gene IDs was selected based on proximity to the CpGs with consistent and nominally significant (p <0.05) estimates in all three models; i.e., 64 CpGs with potential *DNAm* → *Current Smoking* effects ("Nearest Gene" in **Supplementary Table S3**) and 525 CpGs with potential *Current Smoking* → *DNAm* effects ("Nearest Gene" in **Supplementary Table S1**). None of the sites with potential *DNAm* → *Current Smoking* effects are located in the MHC region. For *Current Smoking* → *DNAm* effects, 21 additional sites in the MHC region showed consistent, nominally significant estimates. There was no significant relationship between a CpG site having consistent causal estimates and its being located in the MHC region (Fisher's exact test p-value = 0.5455). However, out of an abundance of caution, the sites located in this region were not included in the enrichment analyses to avoid sites with potentially unreliable results due to its complex LD structure.

As described in the Metascape manuscript [26], the program performed integrated enrichment analyses against multiple reference ontology knowledgebases, including GO processes [27], KEGG pathways [28], canonical pathways [29], and Reactome gene sets [30]. The significant terms with a hypergeometric p-value <0.01 and >1.5-fold enrichment were clustered into a hierarchical tree based on Kappa-statistical similarities among their gene memberships. The tree was then cast into clusters based on a threshold of 0.3 kappa score to obtain enriched, non-redundant ontology terms.

#### eFORGE (experimentally derived Functional element Overlap analysis of ReGions from EWAS)

We performed *eFORGE 2.0* [31–33] analyses of the selected CpG probe IDs with consistent and nominally significant (p <0.05) estimates in either direction (from **Supplementary Tables S1, S3**). Using the web-based tool (https://eforge.altiusinstitute.org/), we examined the overlap between the implicated CpGs and multiple comprehensive reference sets of genomic and epigenomic features that regulate gene expression in different tissues and cell types. The platform was set as “Illumina 450k array”, with default analysis options: proximity = 1kb window, background repetitions = 1000, and significance thresholds of FDR <0.01 (strict) and FDR <0.05 (marginal). Three sets of analyses were performed for each list of probe IDs, selecting the reference data from “Consolidated Roadmap Epigenomics - Chromatin - All 15-state marks”, “Consolidated Roadmap Epigenomics - DHS”, and “Consolidated Roadmap Epigenomics - All H3 marks”.

The eFORGE results include the specific probe IDs overlapping between the input set and the reference sample. We performed iterative follow-up analyses for the CpGs with potential *DNAm → Current Smoking* effects, based on the overlapping probe IDs to examine the specificity of significant (FDR <0.01) enrichment in tissues of interest. Analyses restricted to the 21 CpGs overlapping with enhancers in the fetal brain (**Supplementary Figure S18, Table S12**) showed significant enrichment only for enhancers in the fetal brain samples, suggesting high specificity (**Supplementary Figure S21**). The histone mark analyses also showed enrichment in the fetal brain (though not specific to the brain), wherein all 21 CpGs overlapped with H3K4me1, while a subset of 17 CpGs overlapped with H3K4me3 (**Supplementary Figure S22**). Finally, we performed analyses restricted to these 17 CpGs.

We performed similar follow-up analyses with probe IDs showing overlap with enhancers in the lung (potentially etiologically relevant tissue) and the primary B-cells in cord blood (the tissue type with the most significant enrichment) (from **Supplementary Figure S18, Table S12**).

Enrichment in blood cell types may be influenced by residual cell-composition effects in whole blood analyses [31]. So, we also examined the overlap between the CpGs with potential *DNAm → Current Smoking* effects and the genes implicated in the GWAS of blood cell counts [34] to probe the potential impact of the cell-count GWAS associations on the causal inference and cell-type enrichment. Similar overlap was examined for the subset of CpGs overlapping with enhancers in cord blood primary B cells.

### Supplementary Figures

#### Figure S1

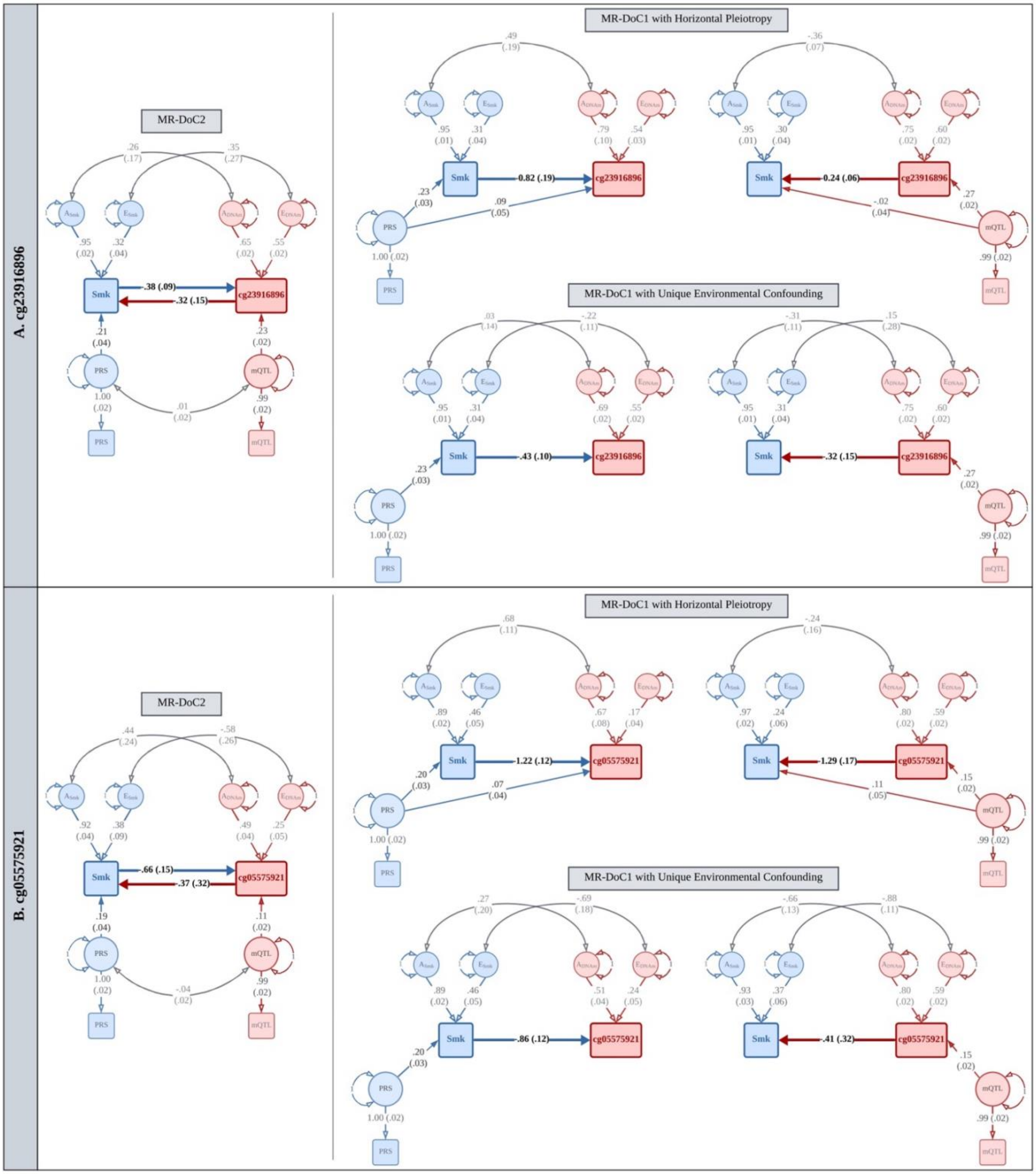

**Illustrative MR-DoC models of causality between current smoking and blood DNAm at (A) cg23916896 and (B) cg05575921 in the AHRR gene.**

*We fitted five MR-DoC models at each CpG: (1) Smoking → DNAm MR-DoC1 with horizontal pleiotropy, (2) Smoking → DNAm MR-DoC1 with unique environmental confounding, (3) DNAm → Smoking MR-DoC1 with horizontal pleiotropy, (4) DNAm → Smoking MR-DoC1 with unique environmental confounding, and (5) bidirectional MR-DoC2. Thus, for each CpG, three causal estimates were obtained in either direction of causation.*

*In the path diagrams, squares/rectangles indicate observed variables, circles indicate latent (unobserved variables), single-headed arrows indicate regression paths, and double-headed curved arrows indicate (co-)variance. The residual variance of smoking status liability is partitioned into additive genetic ($A_{Smk}$) and unique environmental ($E_{Smk}$) components. Likewise, the residual variance of DNAm is partitioned into $A_{DNAm}$ and $E_{DNAm}$. The correlation between $A_{Smk}$ and $A_{DNAm}$ represents the confounding between smoking and DNAm due to latent (unobserved) additive genetic factors, while the correlation between $E_{Smk}$ and $E_{DNAm}$ represents confounding due to latent unique environmental factors. Each model included age and sex as covariates of smoking status (not shown). DNAm $\beta$-values were residualized for standard biological and technical covariates used in EWAS (see Methods). The smoking PRS and the mQTL allelic scores were residualized for standard GWAS covariates, including genetic principal components and genotyping platform. In the path diagrams, the residualized PRS and mQTL allelic scores are regressed on respective latent factors, representing the underlying "true" standardized scores (mean = zero; variance = one). The coefficient of the path from the latent score to the observed score estimates the standard deviation of the observed score.*

*Note. The paths are labeled by the point estimate and its S.E. in parentheses. For better readability, the path diagrams show only the within-individual part of the models fitted to data from twin pairs.*

**Figure S2**

*QQ Plot of MR-DoC1 models (with unique environmental confounding, rE) of Current Smoking → DNAm at 411,169 epigenome-wide CpGs (Bayesian genomic inflation factor, λ = 1.09).*

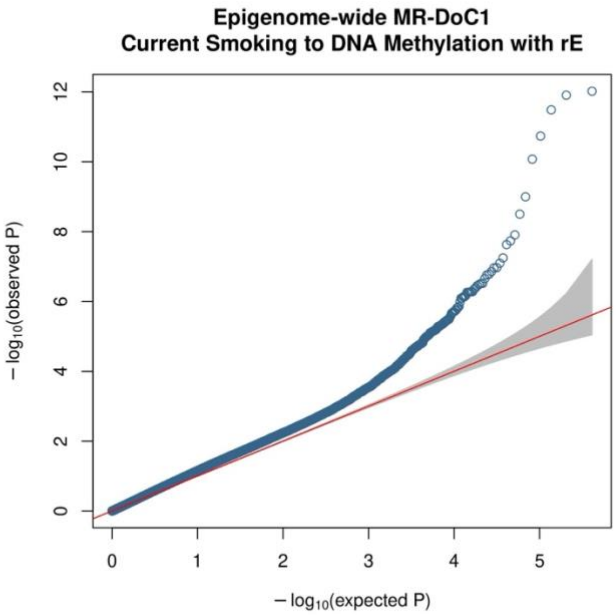

**Figure S3**

*QQ Plot of MR-DoC1 models (with unique environmental confounding, rE) of Current Smoking → DNAm at 16,940 smoking-associated CpGs (Bayesian genomic inflation factor, λ = 1.20).*

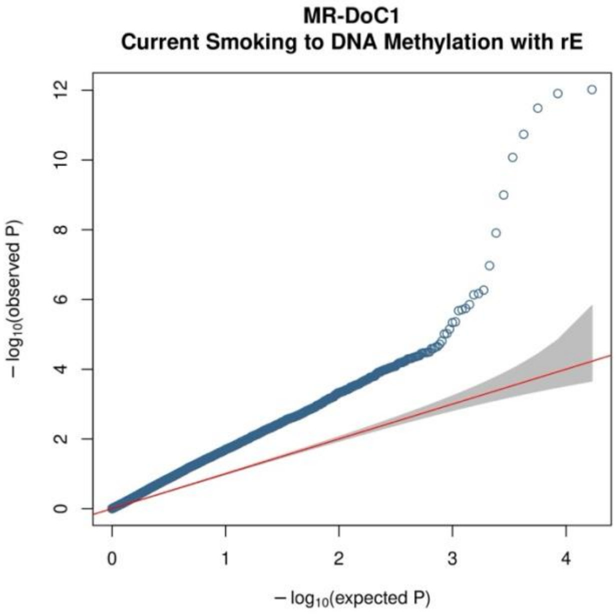

**Figure S4**

*QQ Plot of the Current Smoking → DNAm causal estimates in MR-DoC2 models across 11,124 smoking-associated CpGs (Bayesian genomic inflation factor, λ = 1.20).*

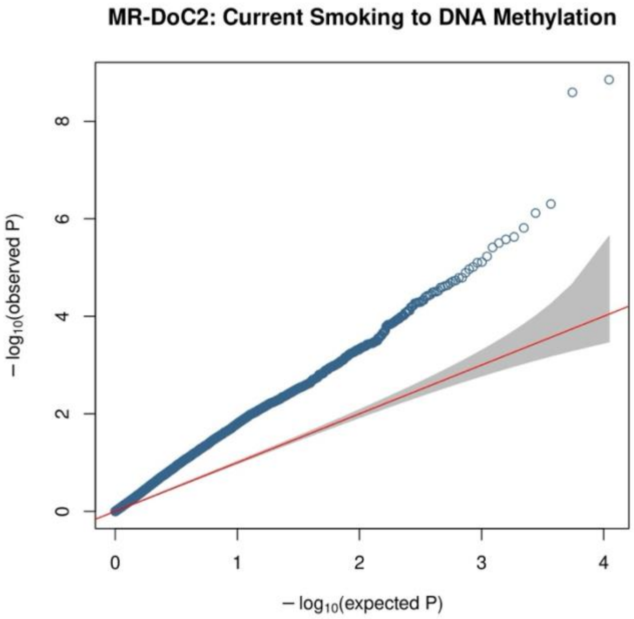

**Figure S5**

*QQ Plot of the DNAm → Current Smoking causal estimates in MR-DoC2 models across 11,124 smoking-associated CpGs (Bayesian genomic inflation factor, λ = 1.01).*

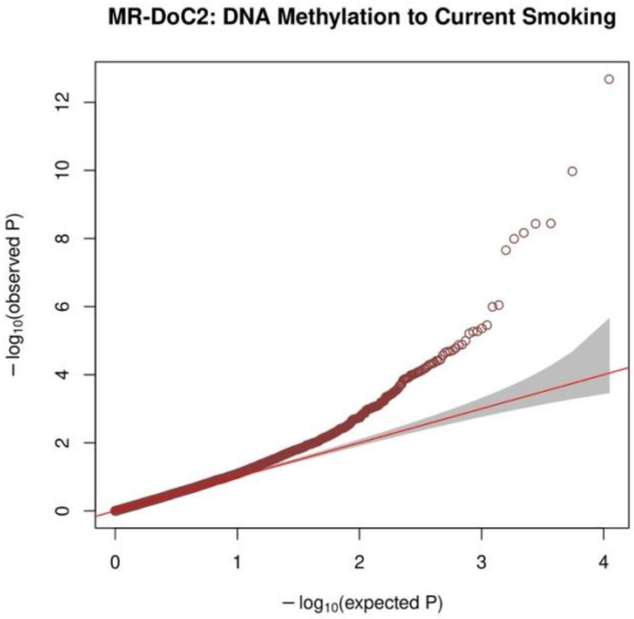

**Figure S6**

*Bidirectional Causal Estimates at the 64 CpGs with Robust Evidence of the Causal Effects of Current Smoking on DNA methylation*

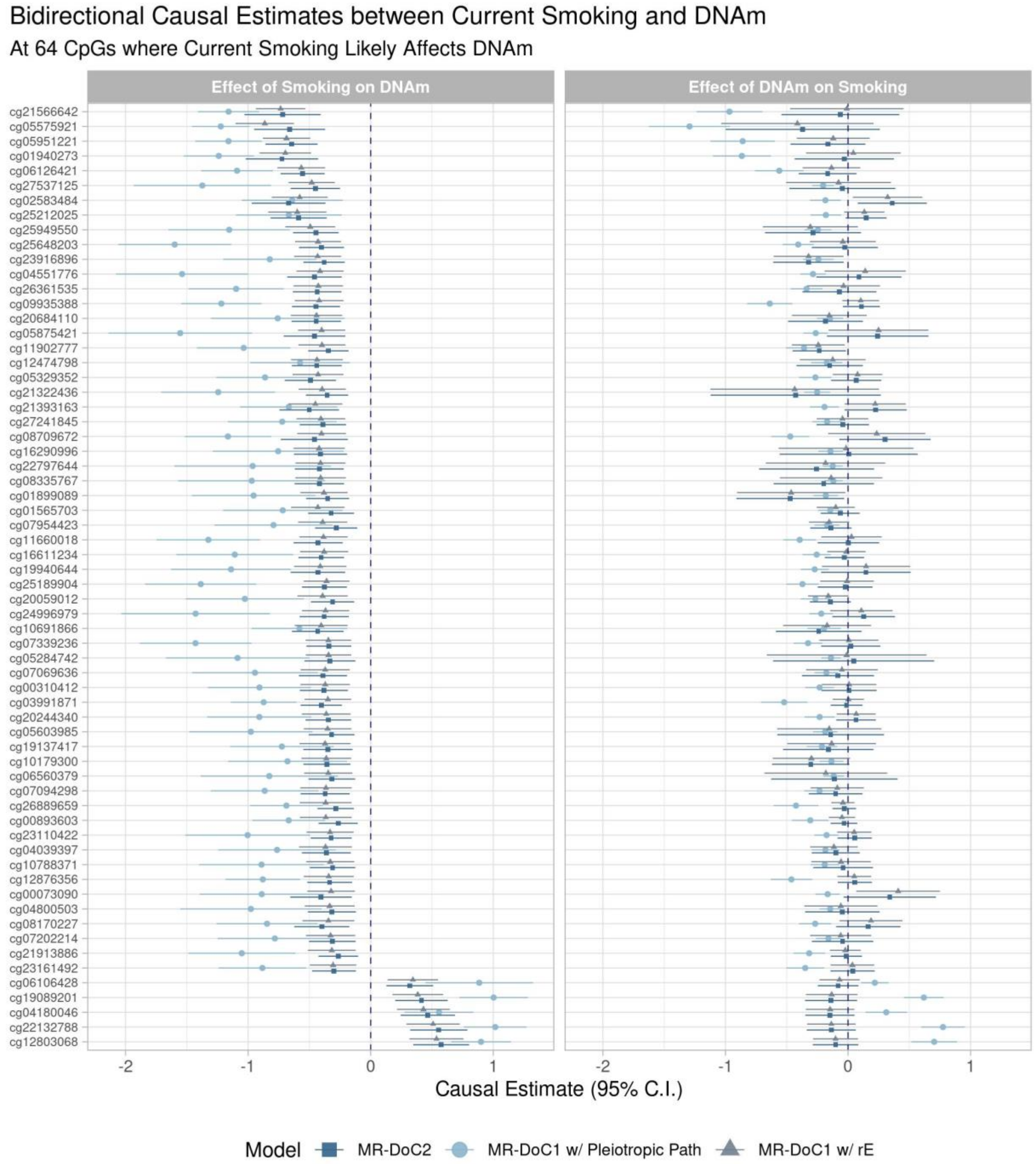

These CpGs did not show robust evidence for the reverse effects of DNAm on current smoking. Please refer to **Supplementary Tables S1** (*Current Smoking* → *DNAm*) and **S2** (*DNAm* → *Current Smoking*) for the corresponding data.

**Figure S6**

*Upset plot of the intersection of CpGs with statistically significant Current Smoking → DNAm effects after Bonferroni correction in each of the three MR-DoC models*

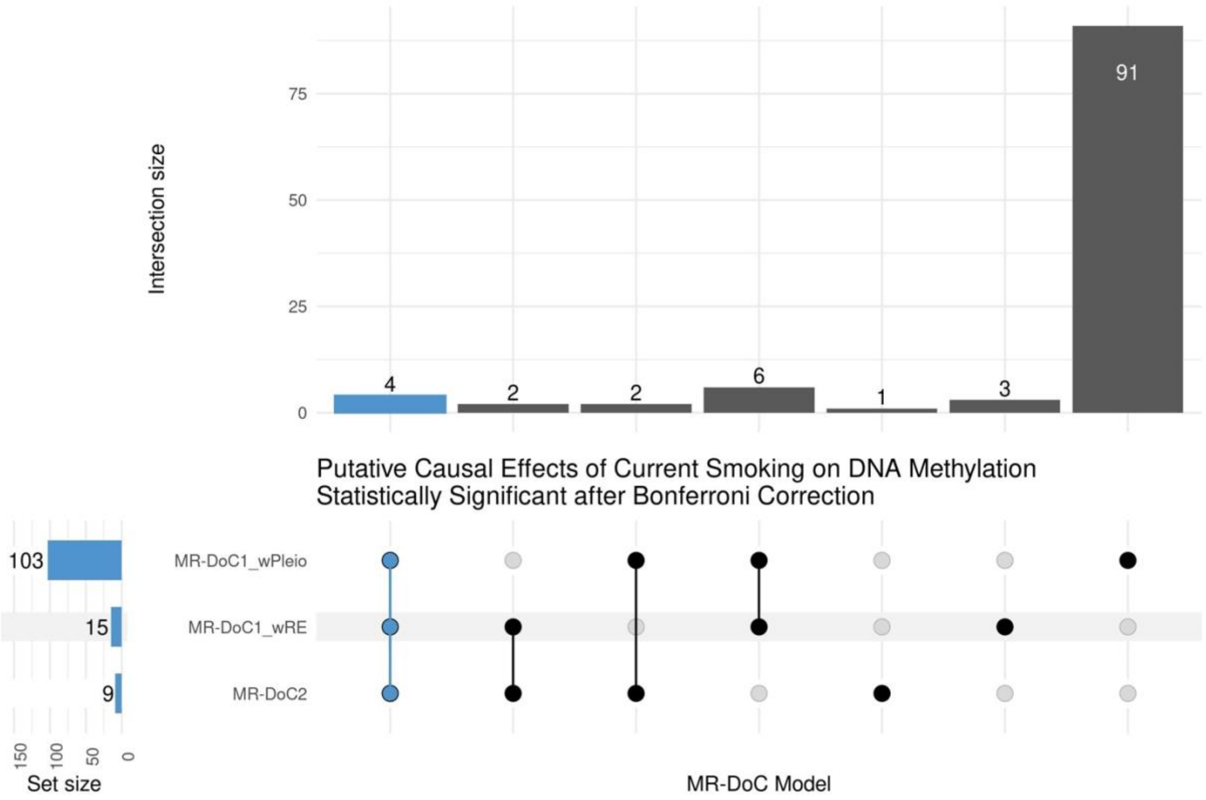

*Note*. Please refer to **Supplementary Table S1** for the corresponding data

**Figure S8**

*Upset plot of the intersection of CpGs with statistically significant DNAm → Current Smoking effects after Bonferroni correction in each of the three MR-DoC models*

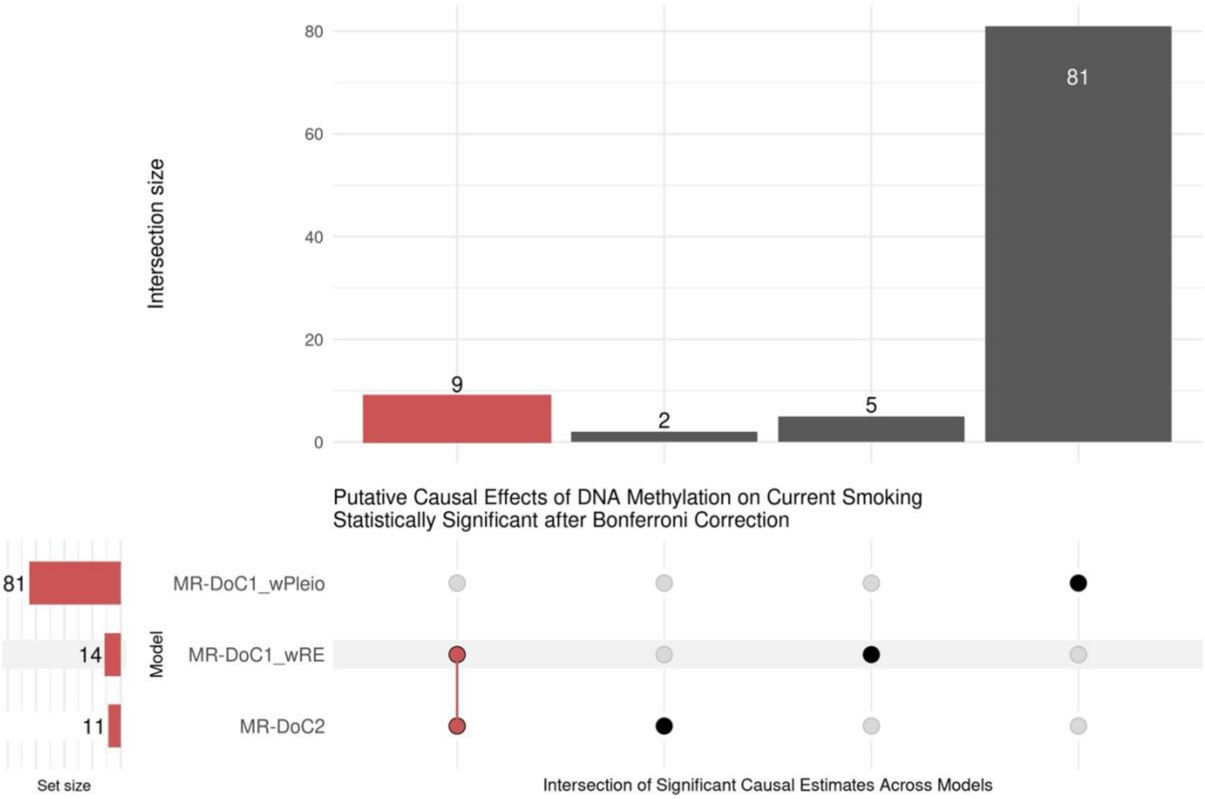

*Note*. Please refer to **Supplementary Table S3** for the corresponding data

**Figure S9**

*19 CpGs with potential bidirectional causal effects between current smoking and DNA methylation*

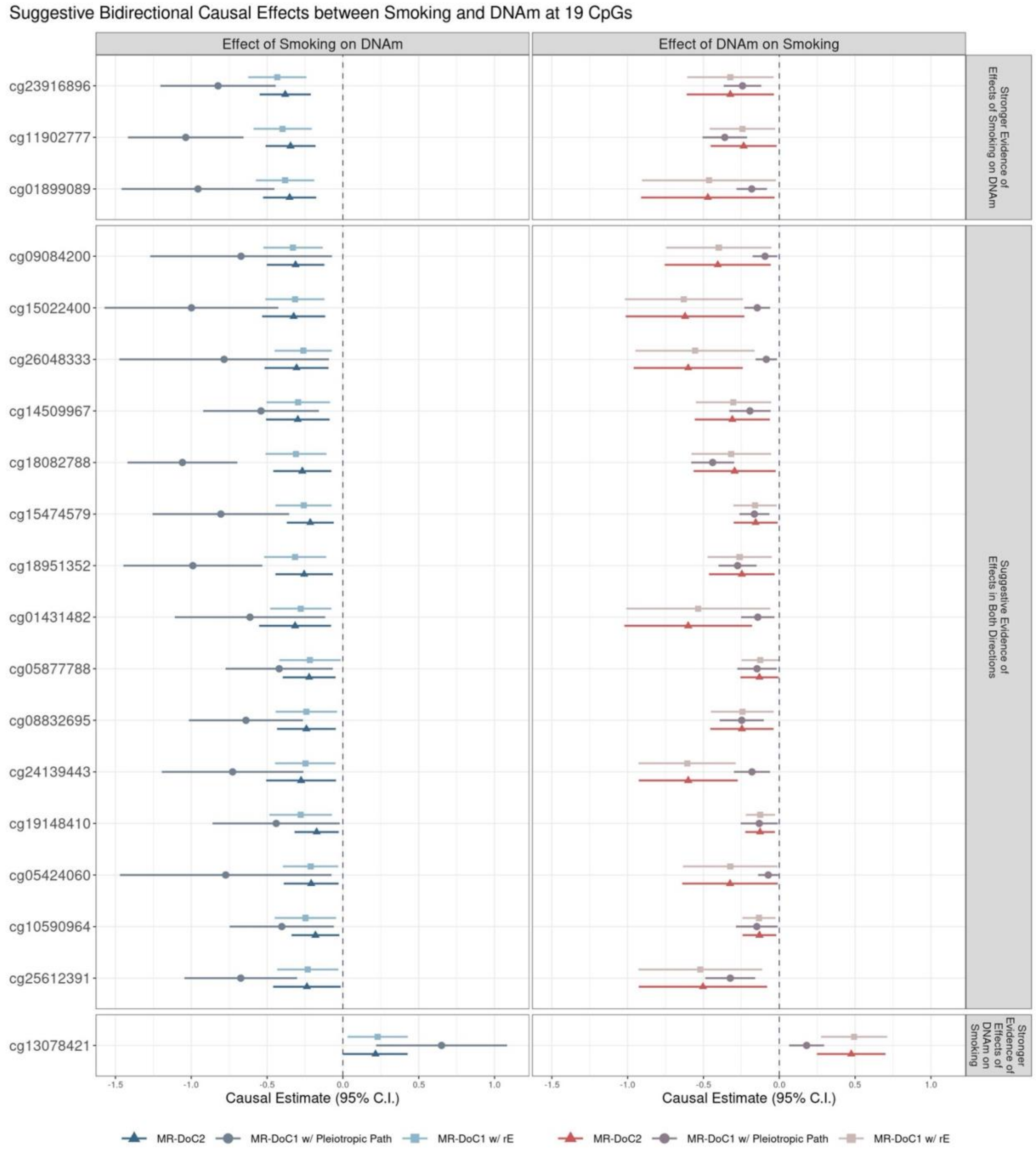

*Note.* Three CpGs had more robust evidence of *Current Smoking* → *DNAm* causal effects than *vice versa*. One CpG had more robust evidence of *DNAm* → *Current Smoking* causal effects than *vice versa*. The rest 15 CpGs had only suggestive evidence (consistent, nominally significant estimates across models) in both directions. Please refer to **Supplementary Tables S1-S4** for the corresponding data.

#### Figure S10

*Top Enriched Ontology Clusters in Metascape's Gene Annotation and Functional Enrichment Analyses of the 525 CpGs (outside the MHC region) with Potential Current Smoking → DNAm effects*

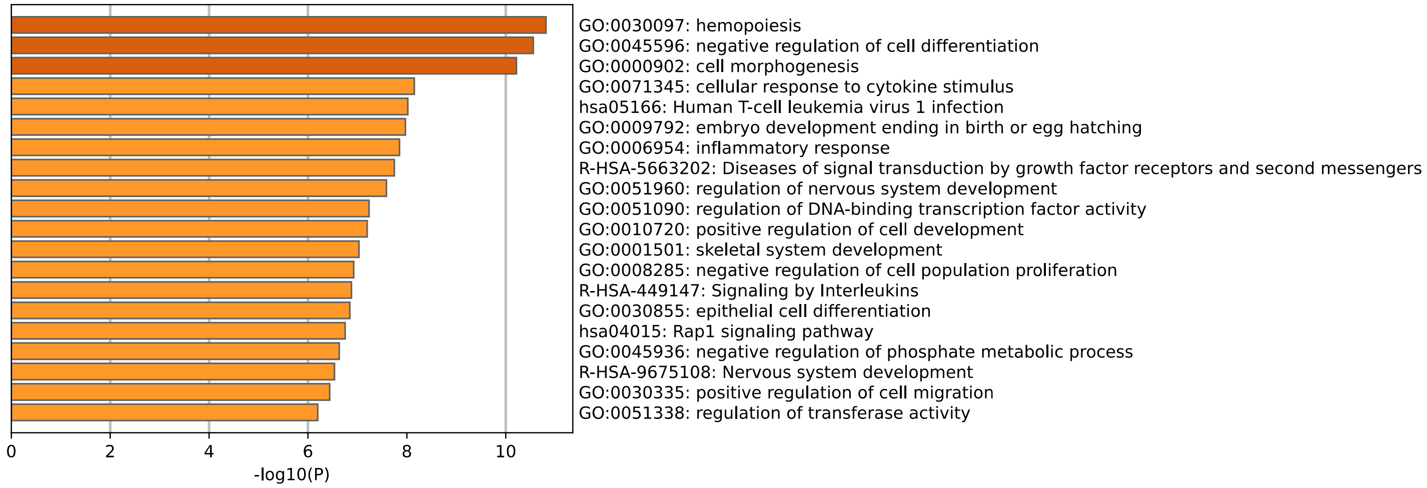

*Note*. The "NearestGene" IDs from *Supplementary Table S1* were used as the input data for Metascape[26]. Please refer to **Supplementary Tables S5-S6** for the corresponding annotation and enrichment results.

As detailed in the Metascape manuscript[26], the program first identified all significant ontology terms, including GO/KEGG terms, canonical pathways, and hallmark gene sets. The significant terms (based on hypergeometric p-value <0.01 and >1.5-fold enrichment) were then clustered into a hierarchical tree based on Kappa-statistical similarities among their gene memberships. The tree was then cast into term clusters based on a threshold of 0.3 kappa score. The enrichment clusters and their underlying terms are marked as "Summary" and "Membership", respectively, under the column *GroupID* in *Supplementary Table S24*. The "Summary" terms provide an overview of enriched, non-redundant ontology terms.

#### Figure S11

*Enrichment Results for Gene-Ontology (GO) Processes in Metascape's Gene Annotation and Functional Enrichment Analyses of the 525 CpGs (outside the MHC region) with Potential Current Smoking → DNAm effects*

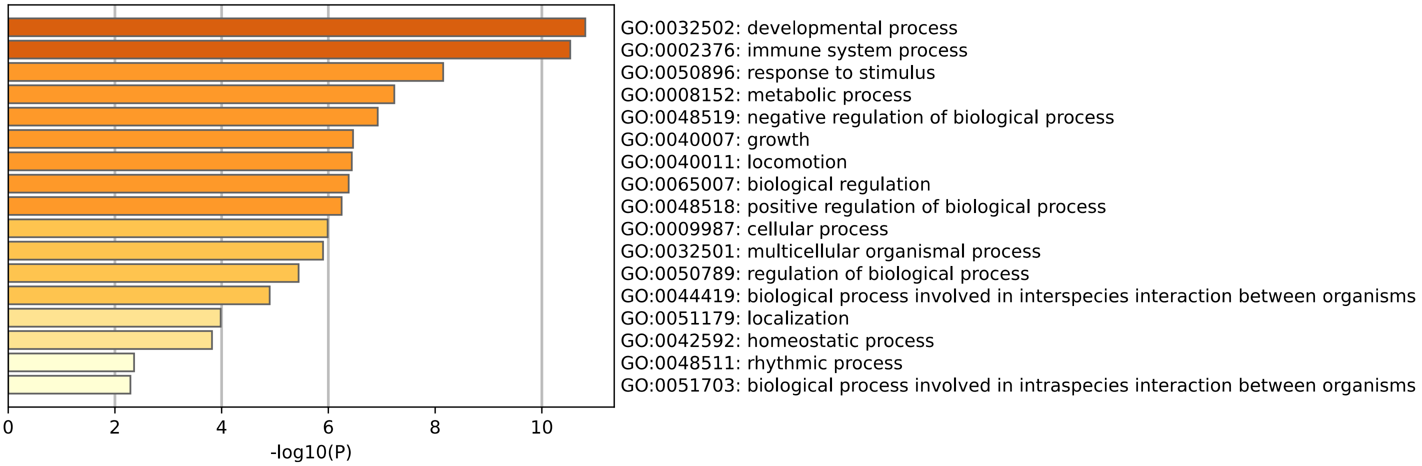

*Note*. The "NearestGene" IDs from *Supplementary Table S1* were used as the input data for Metascape[26]. Please refer to **Supplementary Table S6** for all enrichment results.

#### Figure S12

*Top 100 Ontology Terms in Metascape's Gene Annotation and Functional Enrichment Analyses of the 525 CpGs (outside the MHC region) with Potential Current Smoking → DNAm effects*

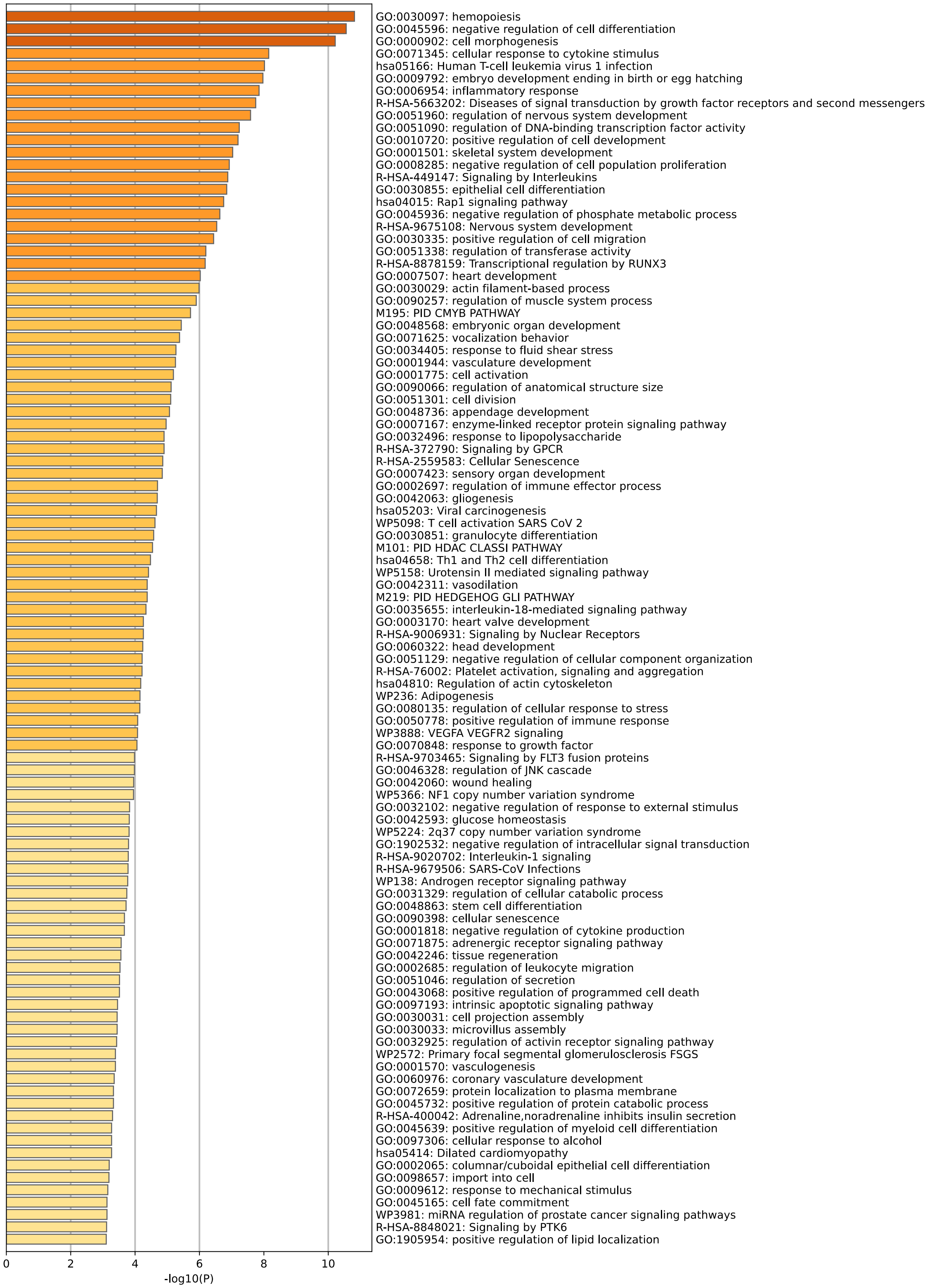

*Note*. The “NearestGene” IDs from *Supplementary Table S1* were used as the input data for Metascape[26]. Please refer to **Supplementary Table S6** for all enrichment results.

#### Figure S13

*eFORGE analyses of overlap between gene-regulatory chromatin states and the 525 CpGs (outside the MHC region) with potential Current Smoking → DNAm effects*

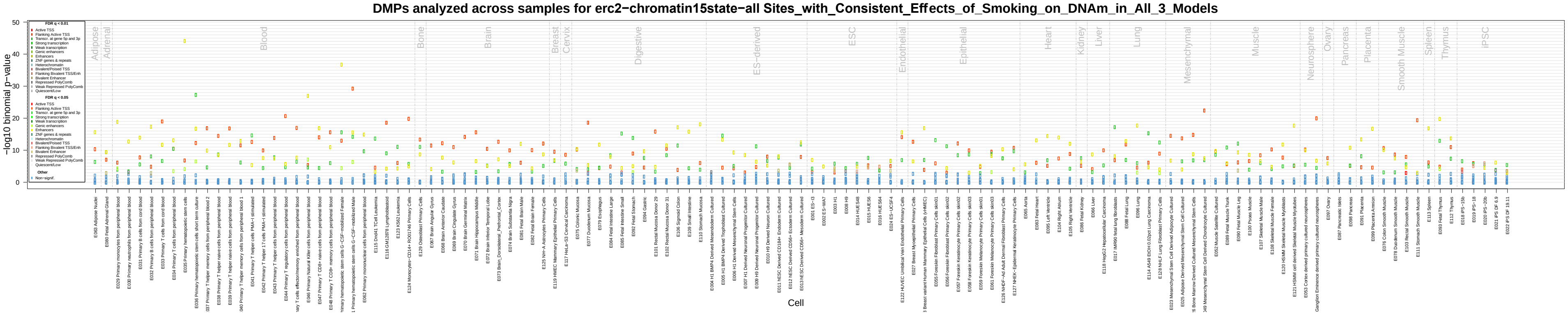

*Note.* Please refer to **Supplementary Table S7** for the corresponding data.

**Figure S14**

*eFORGE analyses of overlap between* histone-mark modifications *and the 525 CpGs (outside the MHC region) with potential Current Smoking → DNAm effects*

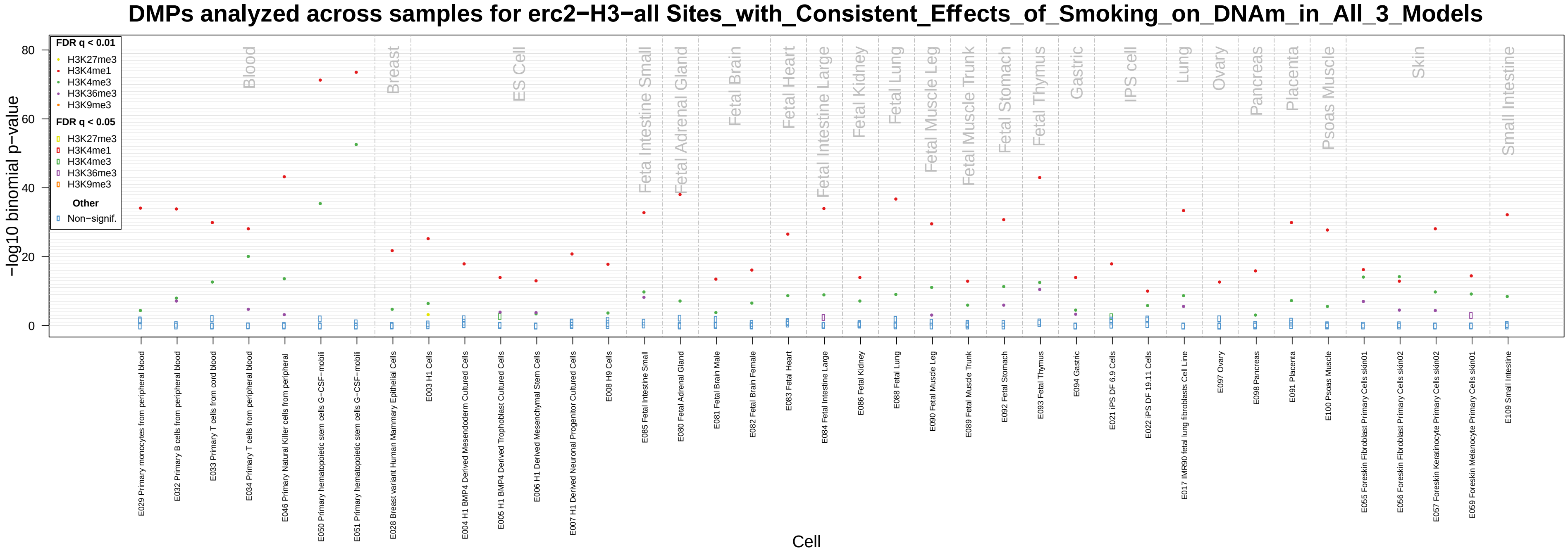

*Note.* Please refer to **Supplementary Table S8** for the corresponding data.

**Figure S15**

*eFORGE analyses of overlap between DNase hypersensitivity (DHS) sites and the 546 CpGs with potential Current Smoking → DNAm effects*

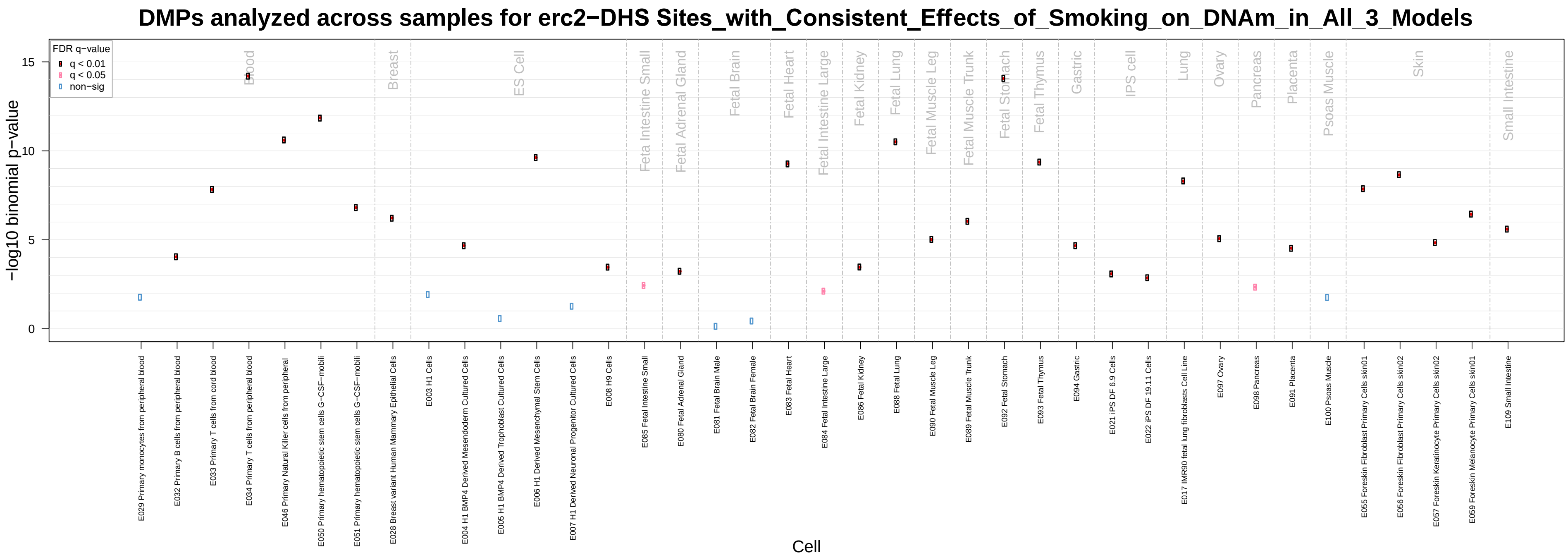

*Note.* Please refer to **Supplementary Table S9** for the corresponding data.

**Figure S16**

*64 CpGs with potential DNAm → Current Smoking effects, based on consistent, nominally significant estimates across models*

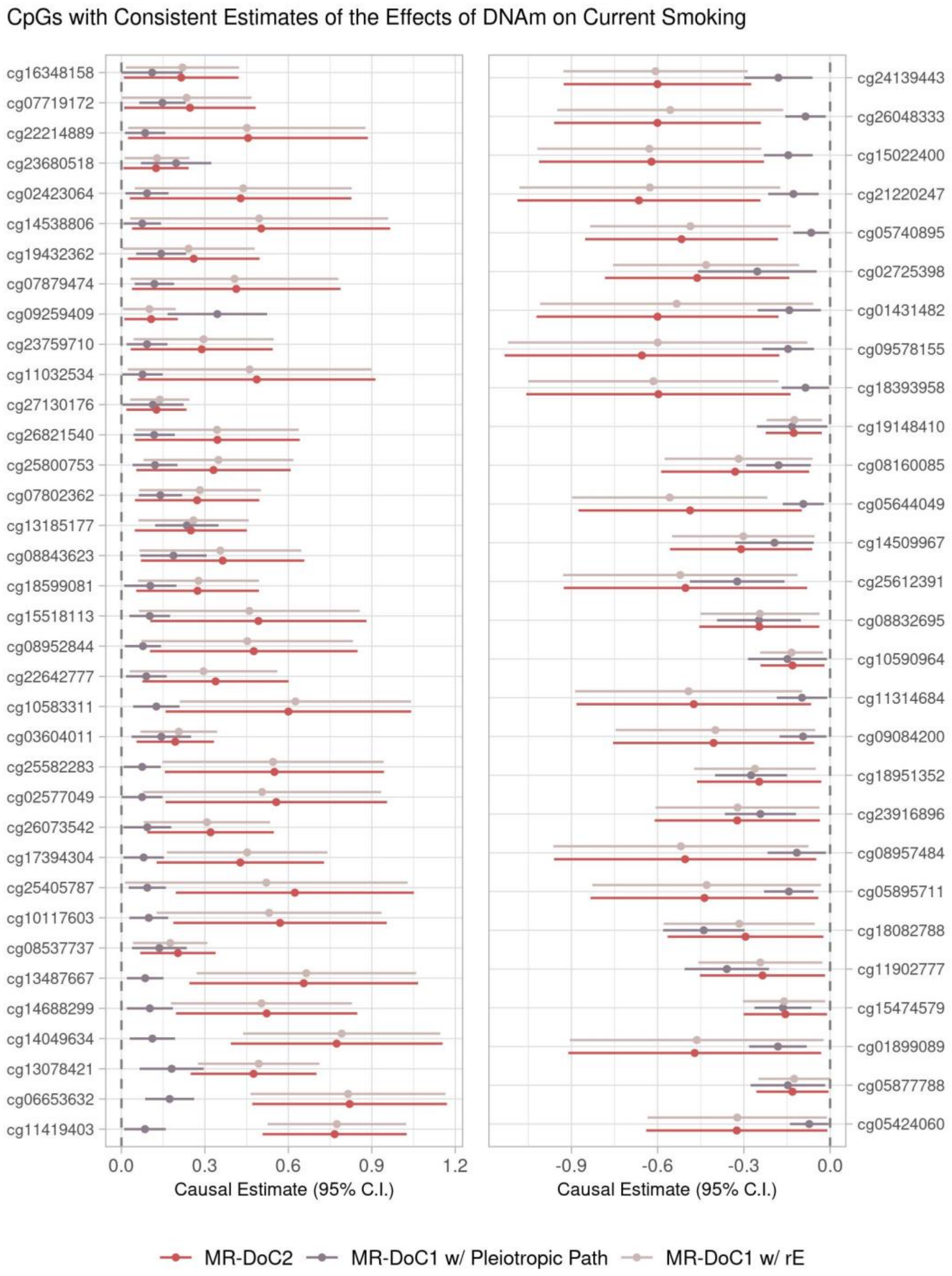

*Note*. These CpGs were used for the follow-up enrichment analyses with eFORGE[31] and Metascape[26]. None of these sites are in the MHC region. Please refer to **Supplementary Table S3** for the corresponding data.

#### Figure S17

*Top Ontology Clusters in Metascape's Gene Annotation and Functional Enrichment Analyses of the 64 CpGs with Potential DNAm → Current Smoking effects*

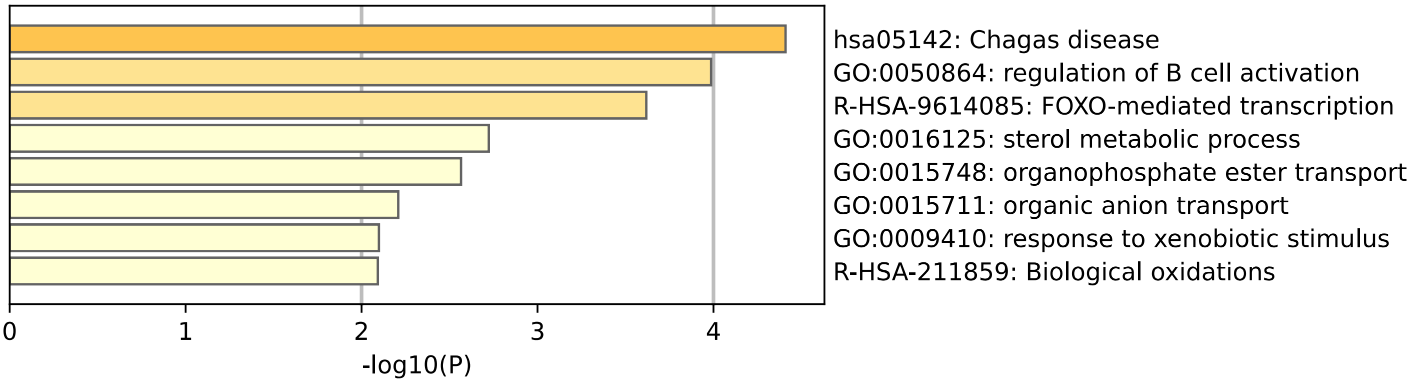

*Note*. The "NearestGene" IDs from *Supplementary Table S3* were used as the input data for Metascape[26]. None of the ontology terms were significant after multiple-testing correction. Please refer to **Supplementary Tables S10 and S11** for all annotation and enrichment results.

#### Figure S18

*Enrichment Results for Gene-Ontology (GO) Processes in Metascape's Gene Annotation and Functional Enrichment Analyses of the 64 CpGs with Potential DNAm → Current Smoking effects*

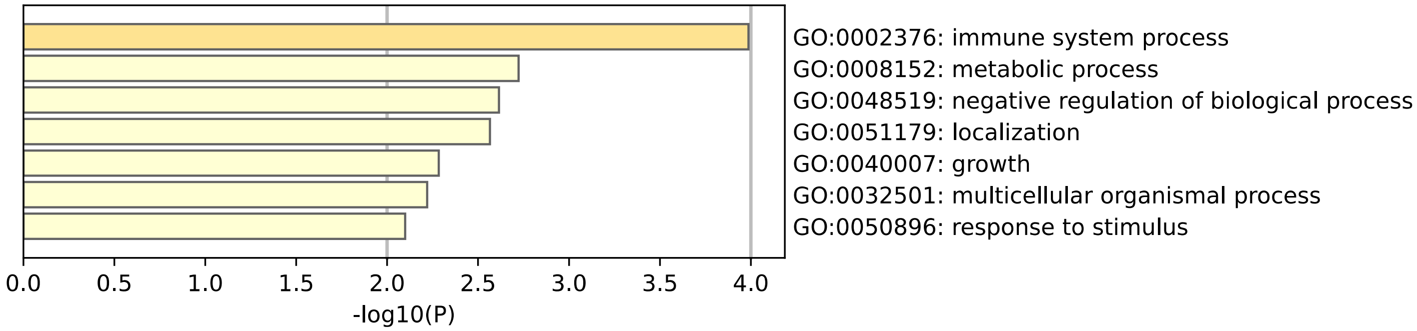

*Note*. The "NearestGene" IDs from *Supplementary Table S3* were used as the input data for Metascape[26]. None of the ontology terms were significant after multiple-testing correction. Please refer to **Supplementary Table S11** for all enrichment results.

#### Figure S19

*eFORGE analyses of overlap between gene-regulatory chromatin states and the 64 CpGs with potential DNAm → Current Smoking effects*

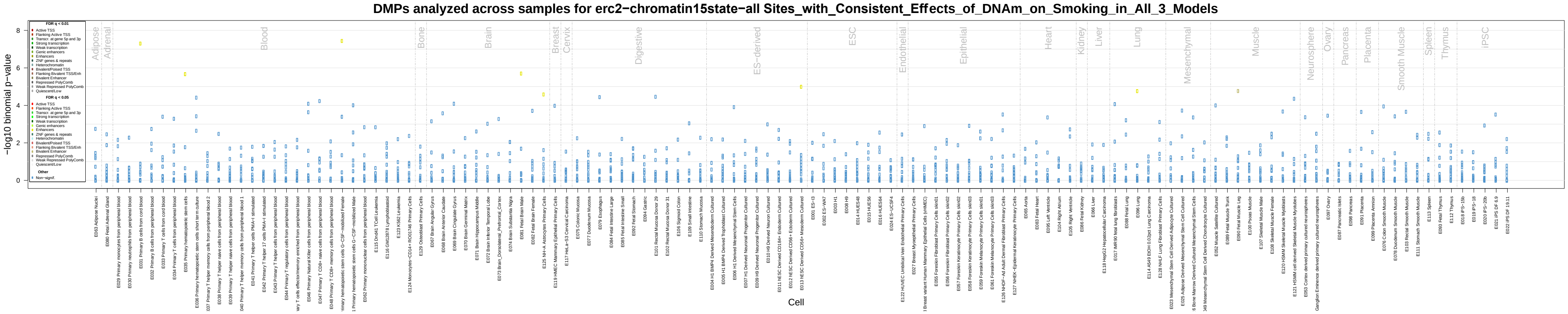

*Note.* Please refer to **Supplementary Table S12** for the corresponding data.

**Figure S20**

*eFORGE analyses of overlap between* histone-mark modifications *and the 64 CpGs with potential DNAm* → *Current Smoking effects*

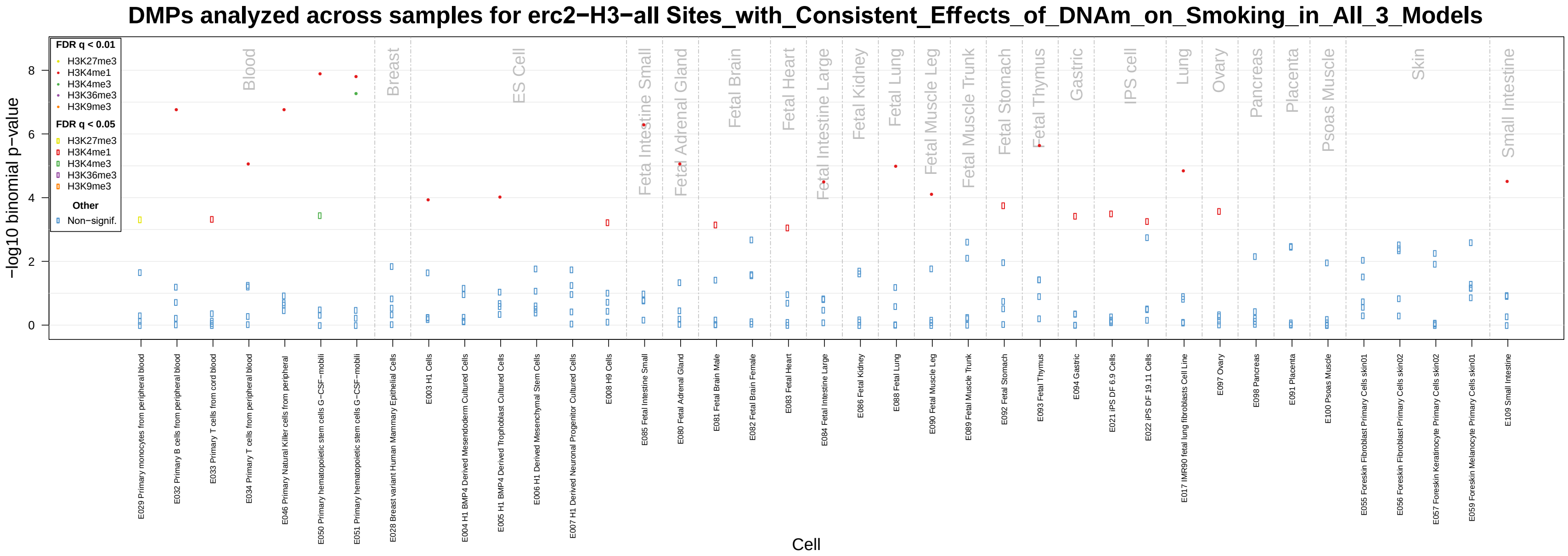

*Note.* Please refer to **Supplementary Table S13** for the corresponding data.

**Figure S21**

*eFORGE analyses of overlap between DNase hypersensitivity (DHS) sites and the 64 CpGs with potential DNAm → Current Smoking effects*

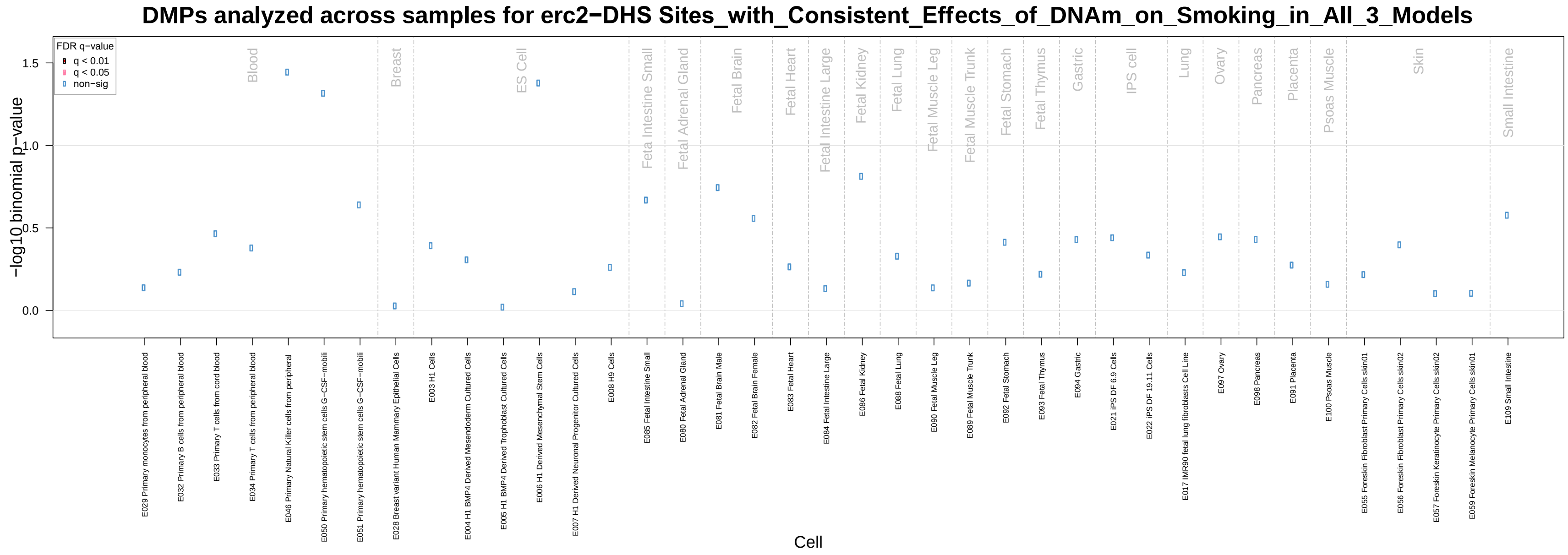

*Note.* Please refer to **Supplementary Table S14** for the corresponding data.

#### Figure S22

*Follow-up eFORGE analyses of overlap between gene-regulatory chromatin states and the 21 CpGs enriched for overlap with Enhancers in the "Fetal Brain Male" sample in Figure S19/Table S5*

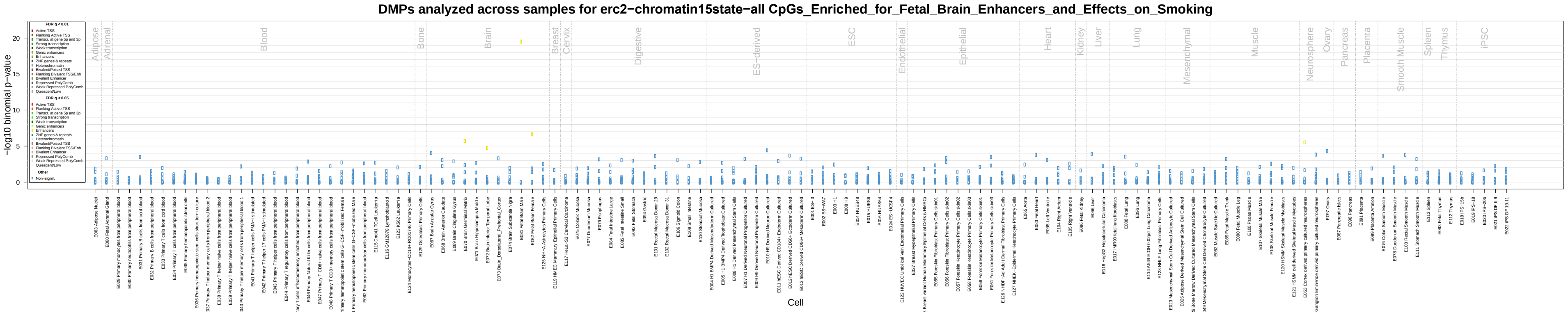

*Note.* Please refer to **Supplementary Table S15** for the corresponding data.

**Figure S23**

*Follow-up eFORGE analyses of overlap between* histone-mark modifications *and the 21 CpGs enriched for overlap with Enhancers in the "Fetal Brain Male" sample in Figure S19/Table S5*

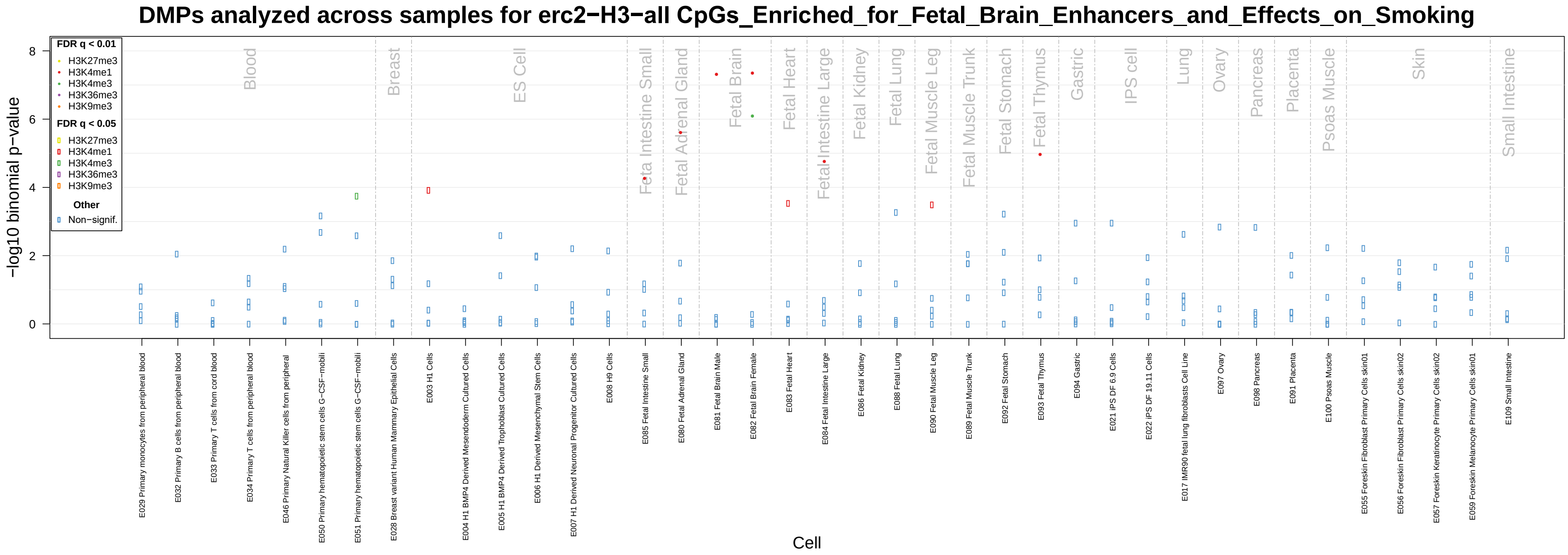

*Note.* Please refer to **Supplementary Table S16** for the corresponding data.

#### Figure S24

*Follow-up eFORGE analyses of overlap between DNase hypersensitivity (DHS) sites and the 21 CpGs enriched for overlap with Enhancers in the "Fetal Brain Male" sample in Figure S19/Table S5*

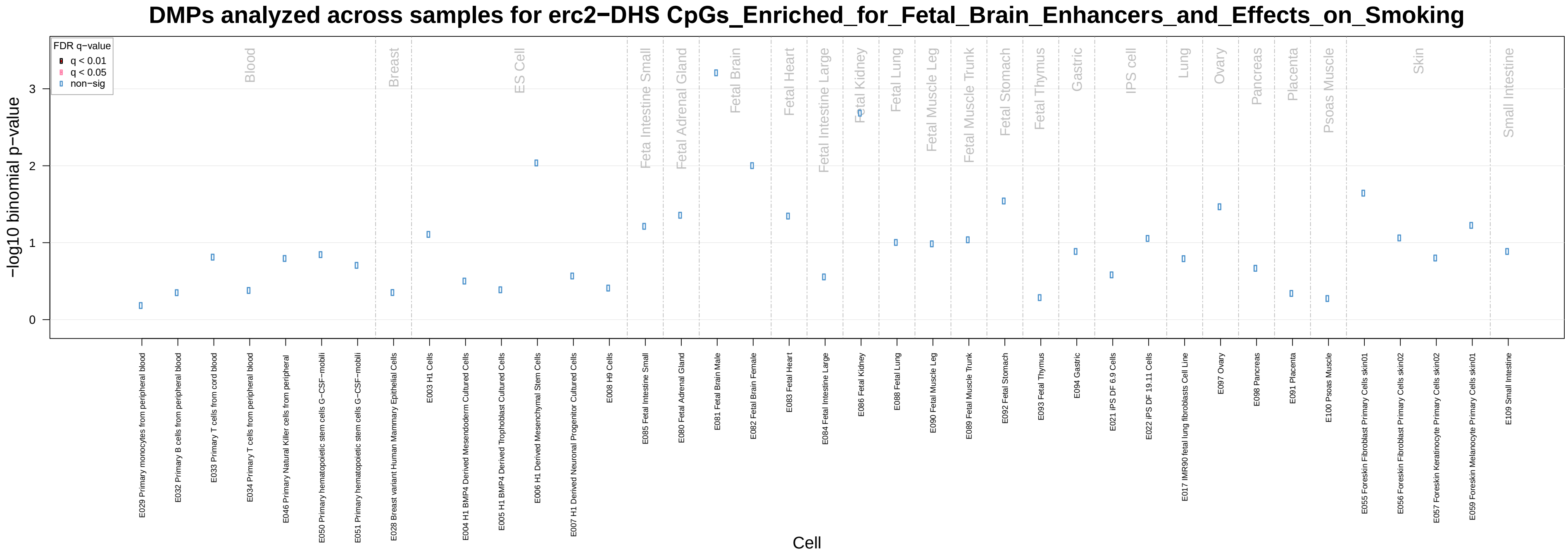

*Note.* Please refer to **Supplementary Table S17** for the corresponding data.

#### Figure S25

*Follow-up eFORGE analyses of overlap between DNase hypersensitivity (DHS) sites and the 17 CpGs enriched for overlap with H3K4me3 modifications in the "Fetal Brain Female" samples in Supplementary Figure S23/Table S9*

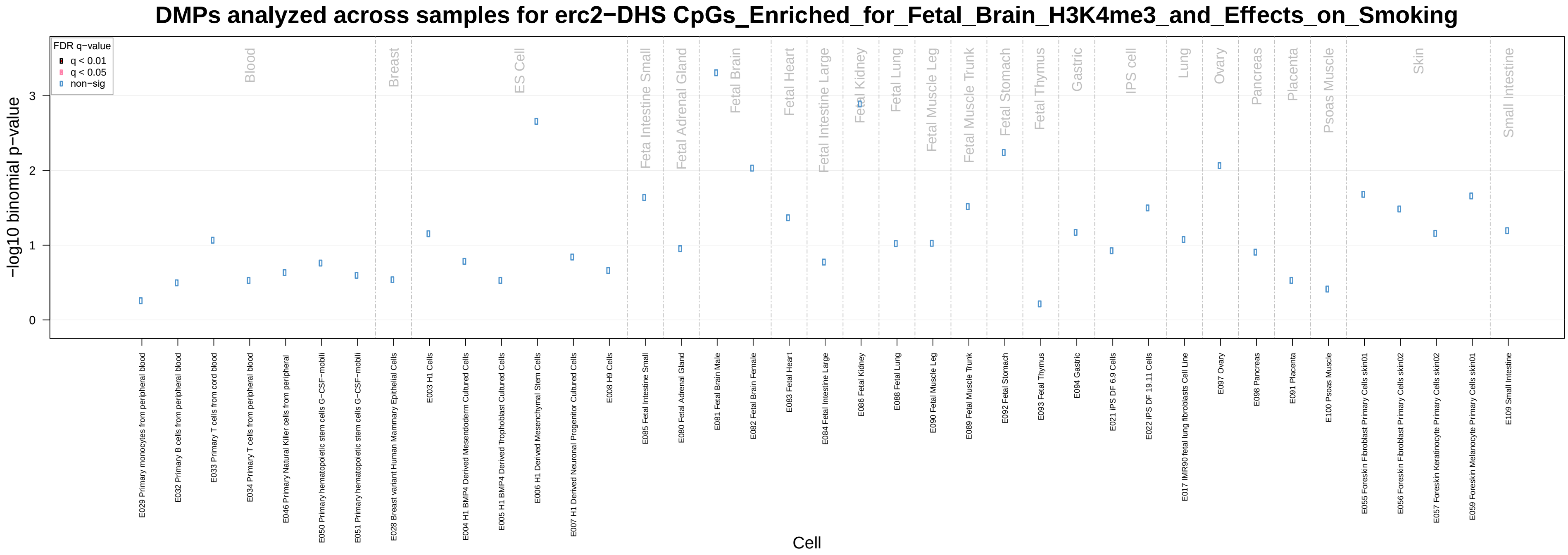

*Note.* Please refer to **Supplementary Table S20** for the corresponding data.

**Figure S26**

*Estimated DNAm → Current Smoking effects at the 17 CpGs showing highly specific enrichment for overlap with gene-regulatory elements in the brain in Figure 5*

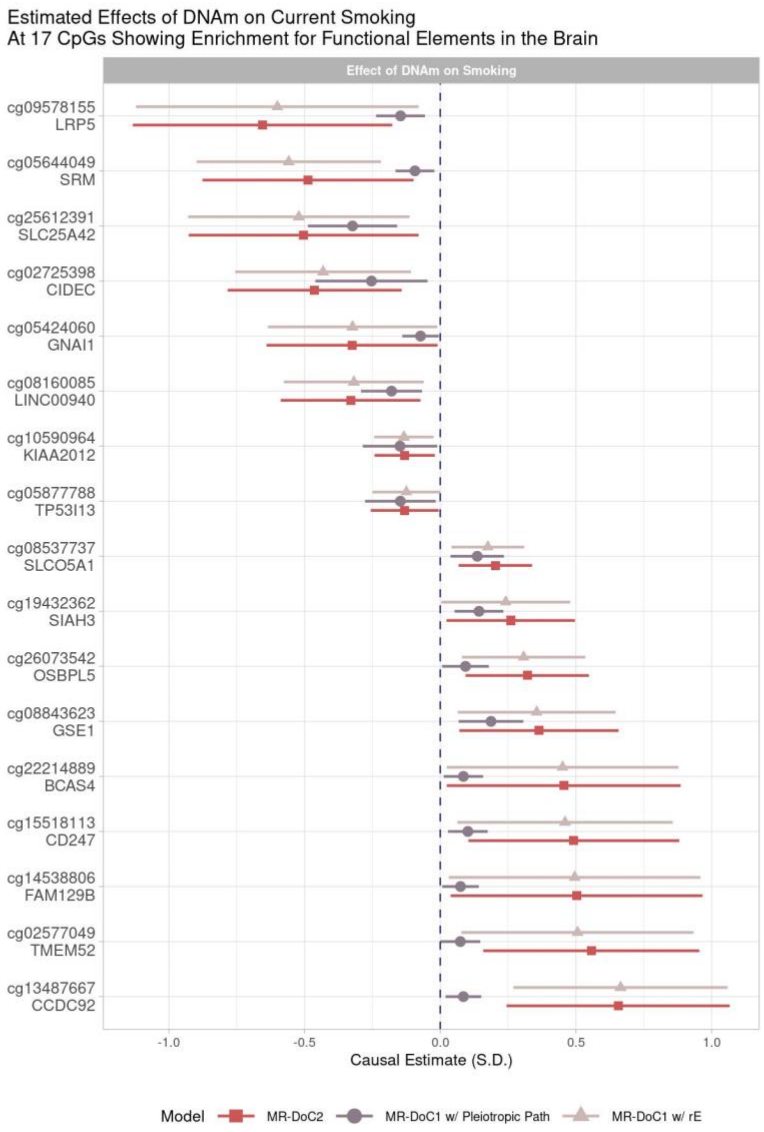

*Note.* The Y-axis shows the probe ID and the "Nearest Gene". For the corresponding data, please refer to **Supplementary Table S3**.

**Figure S27**

*eFORGE analyses of overlap between gene-regulatory chromatin states and the 18 CpGs underlying the enriched overlap with Enhancers in the "Lung" sample in Figure S19/Table S5*

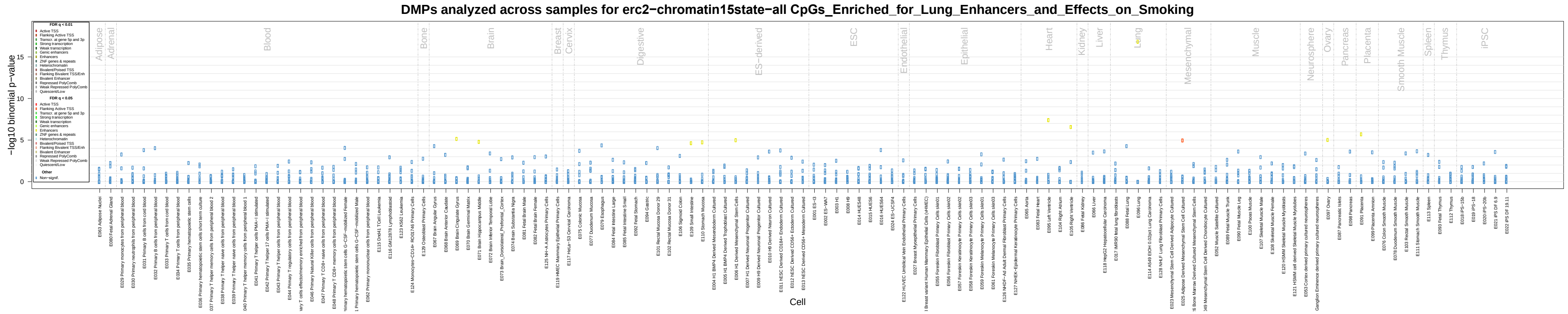

*Note.* Please refer to **Supplementary Table S21** for the corresponding data.

**Figure S28**

*eFORGE analyses of overlap between histone-mark modifications and the 18 CpGs underlying the enriched overlap with Enhancers in the "Lung" sample in Figure S19/Table S5*

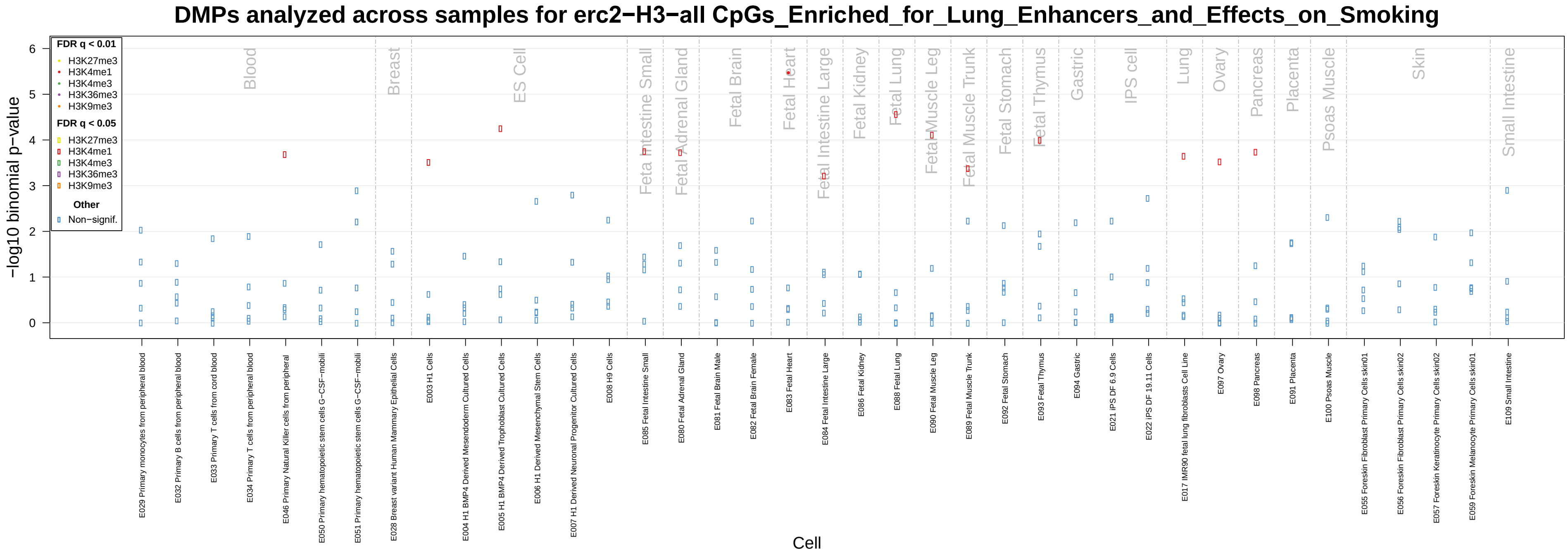

*Note.* Please refer to **Supplementary Table S22** for the corresponding data.

**Figure S29**

*eFORGE analyses of overlap between DNase hypersensitivity (DHS) sites and the 18 CpGs underlying the enriched overlap with Enhancers in the "Lung" sample in Figure S19/Table S5*

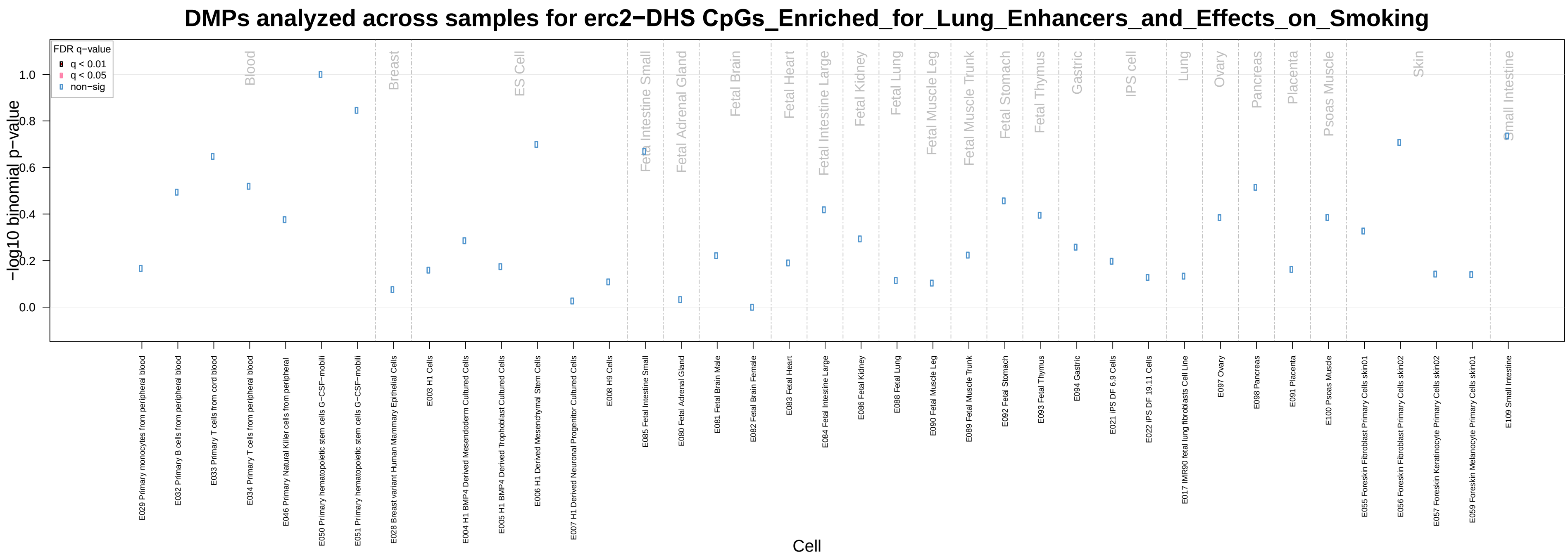

*Note.* Please refer to **Supplementary Table S23** for the corresponding data.

#### Figure S30

*eFORGE analyses of overlap between gene-regulatory chromatin states and the 18 CpGs underlying the enriched overlap with Enhancers in the "Primary B cells from cord blood" sample in Figure S19/Table S5*

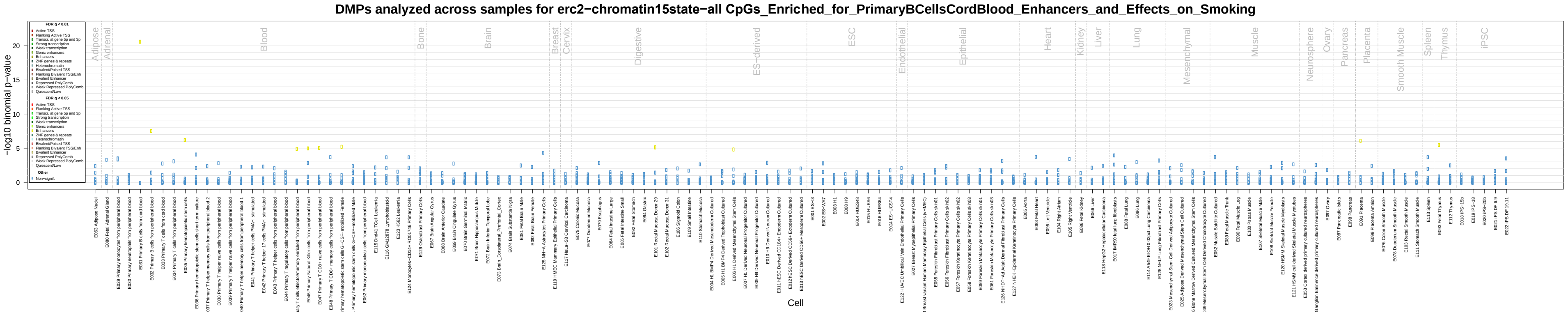

*Note.* Please refer to **Supplementary Table S24** for the corresponding data.

**Figure S31**

*eFORGE analyses of overlap between histone-mark modifications and the 18 CpGs underlying the enriched overlap with Enhancers in the "Primary B cells from cord blood" sample in Figure S19/Table S5*

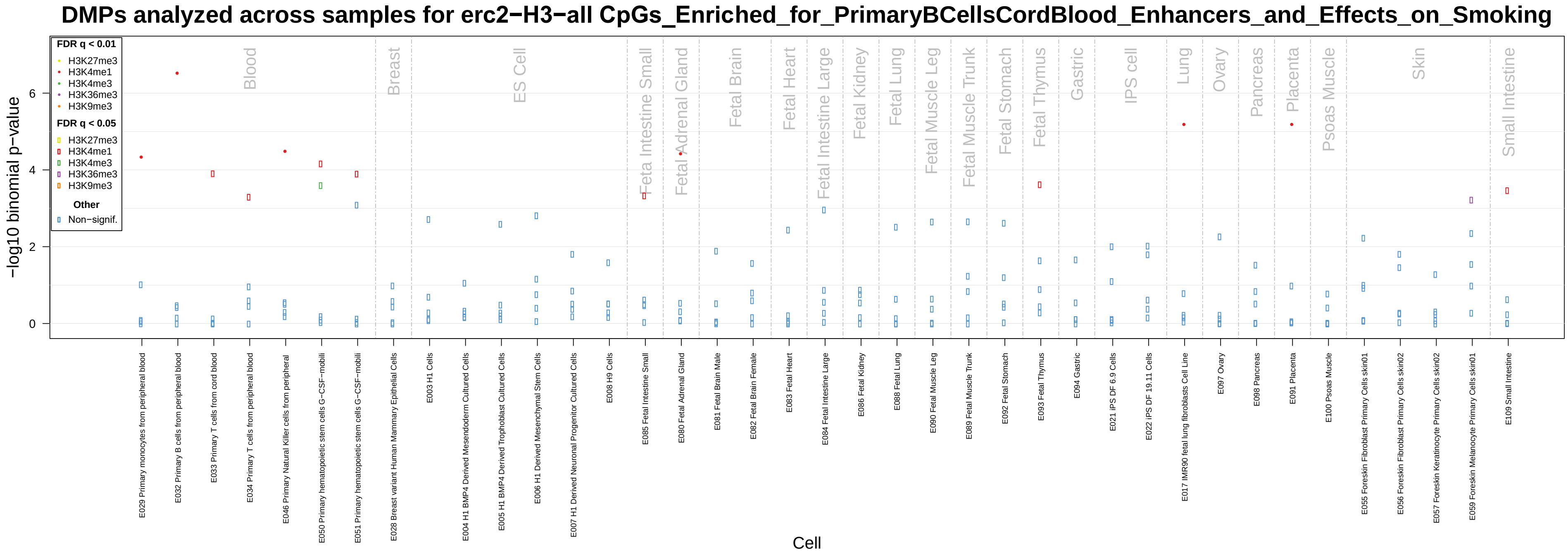

*Note.* Please refer to **Supplementary Table S25** for the corresponding data.

**Figure S32**

*eFORGE analyses of overlap between DNase hypersensitivity (DHS) sites and the 18 CpGs underlying the enriched overlap with Enhancers in the "Primary B cells from cord blood" sample in Figure S19/Table S5*

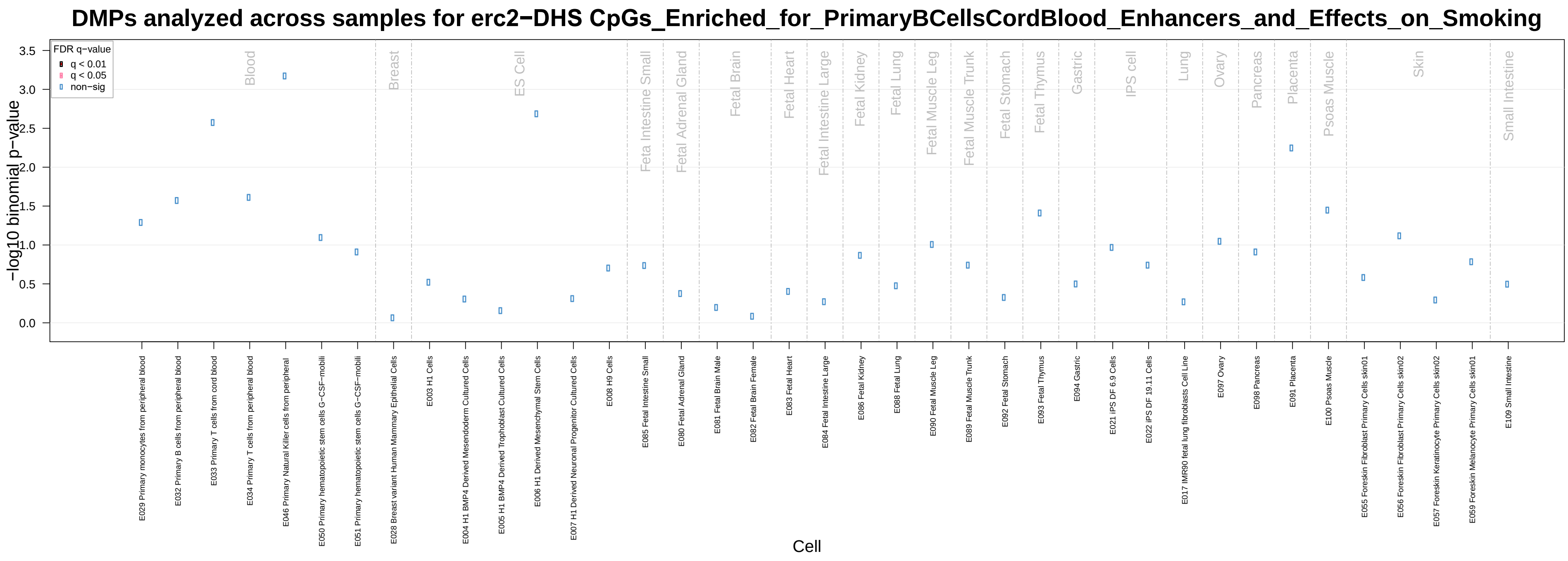

*Note.* Please refer to **Supplementary Table S26** for the corresponding data.

**Figure S33**

*Upset plot of the intersection of CpGs with statistically significant (FDR <0.05) Former Smoking → DNAm effects in each of the three MR-DoC models*

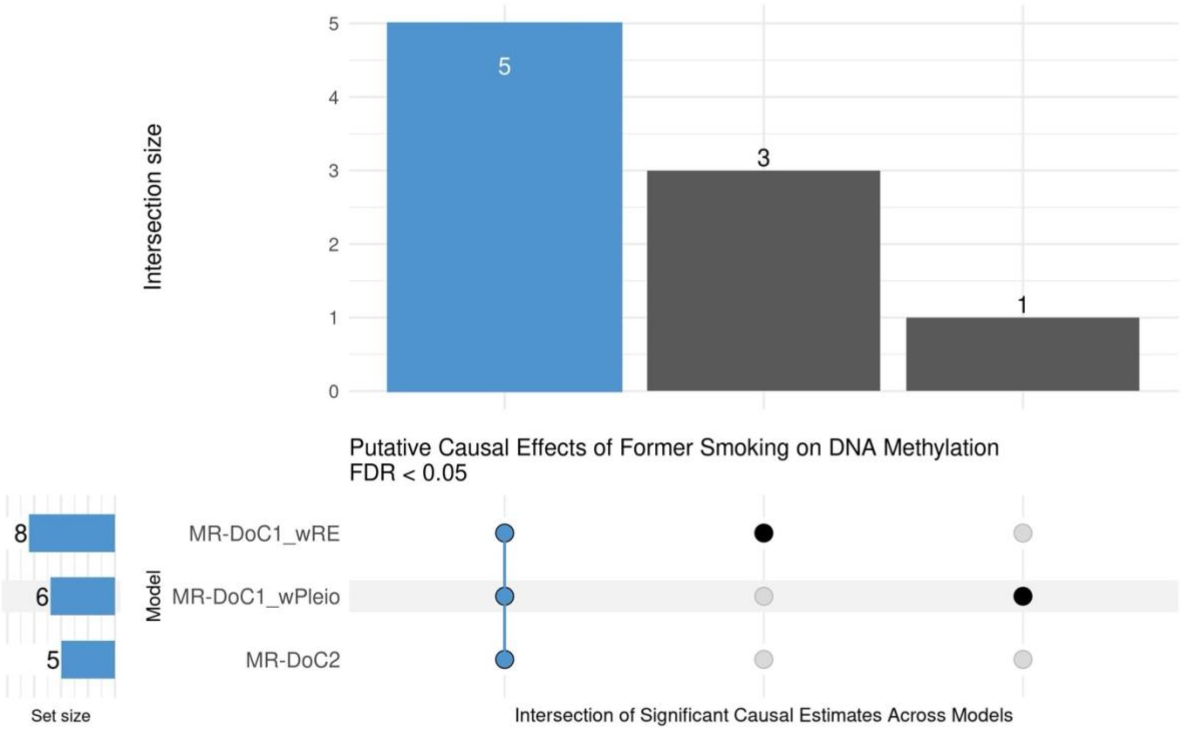

*Note.* Please refer to **Supplementary Table S27** for the corresponding data.

**Figure S34**

*Upset plot of the intersection of CpGs with statistically significant (FDR <0.05) DNAm → Former Smoking effects in each of the three MR-DoC models*

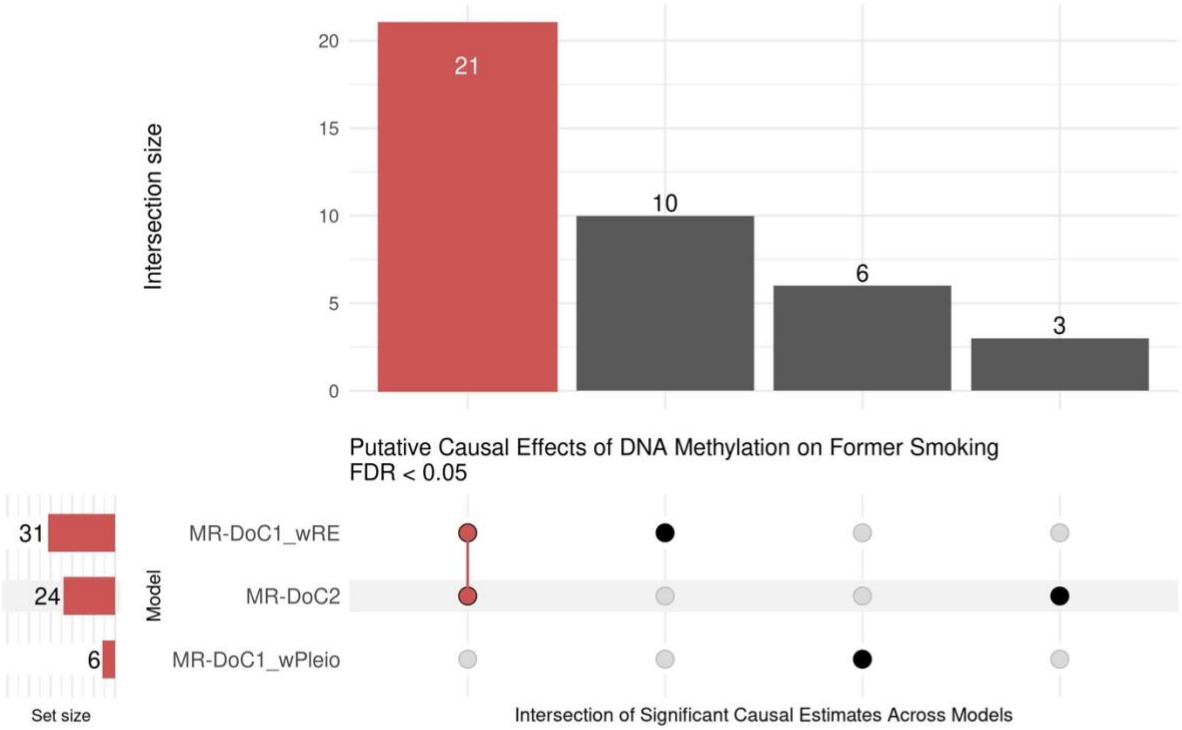

*Note.* Please refer to **Supplementary Table S29** for the corresponding data.

**Figure S35**

*Estimated DNAm → Former Smoking effects at the two former-smoking-associated CpGs that showed robust evidence of DNAm → Current Smoking effects*

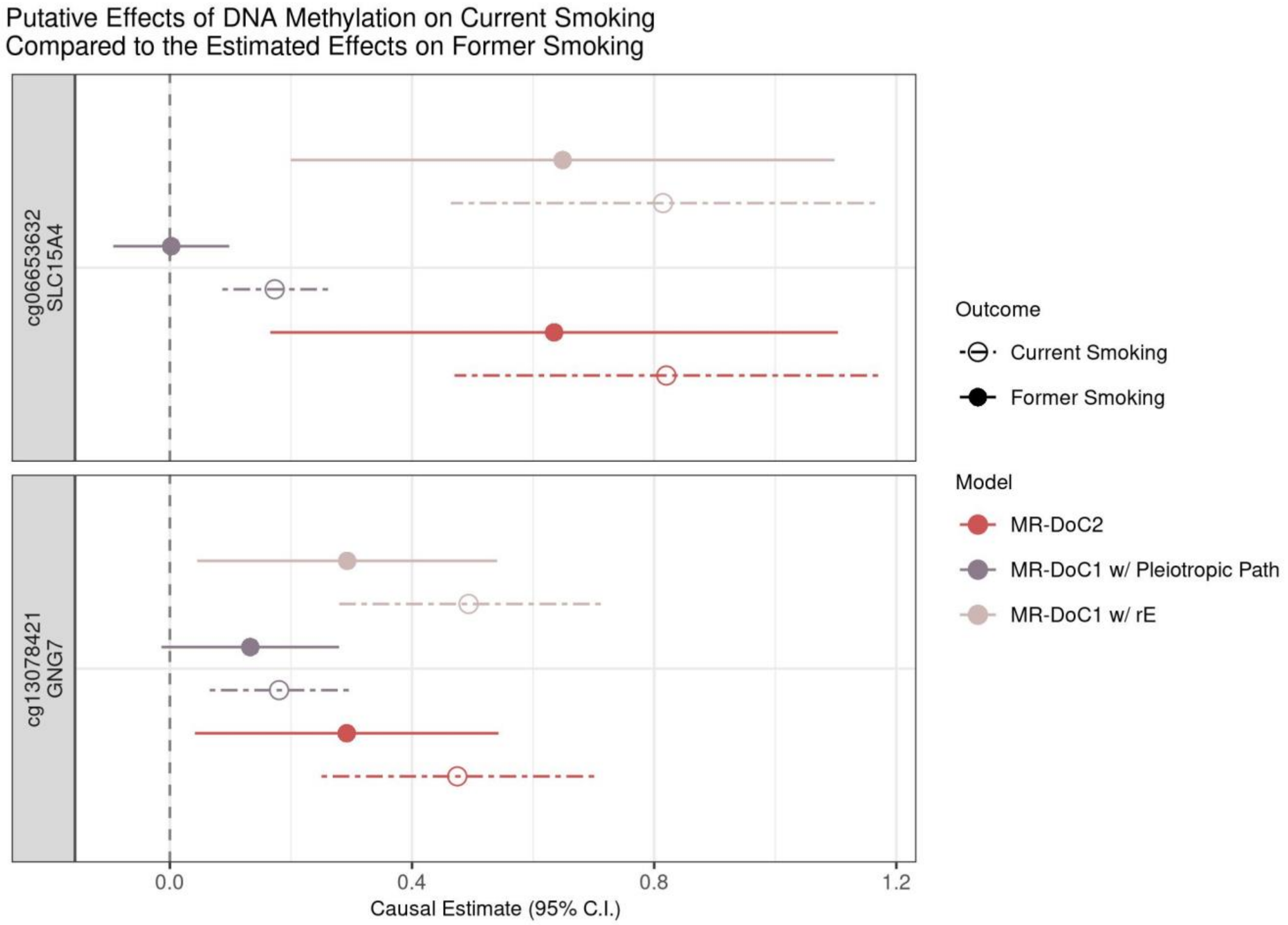

*Note.* Please refer to **Supplementary Tables S3 and S29** for the corresponding data.

**Figure S36**

*Prior EWAS association statistics of smoking-associated CpGs stratified by whether the CpG was identified as having an mQTL allelic score with F-statistic >10 in the current study*

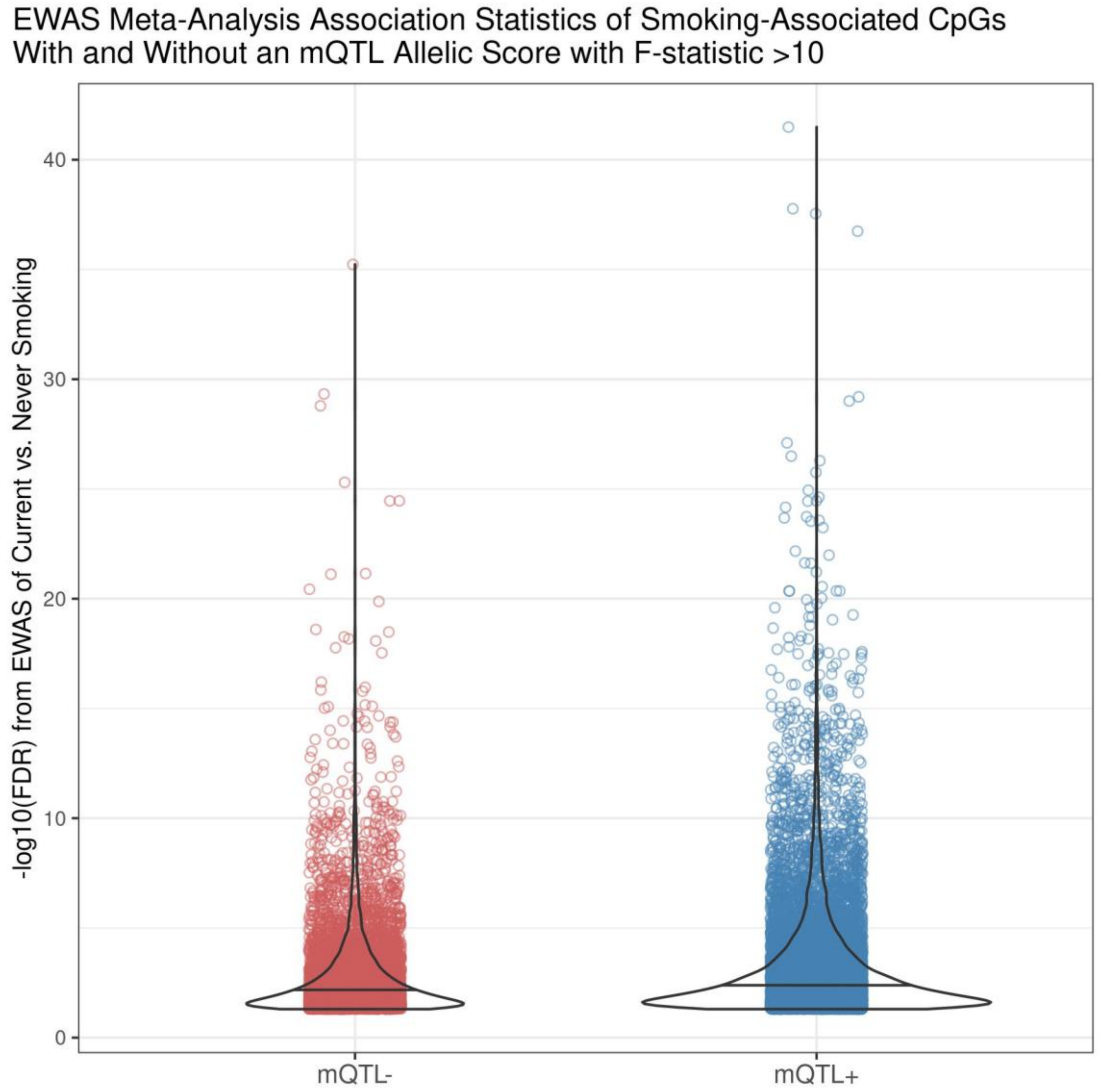

*On the X-axis, "mQTL-" indicates the CpGs without an mQTL allelic score with F >10 (5,816 CpGs), and "mQTL+" indicates the CpGs with an mQTL allelic score with F >10 (11,124 CpGs). The Y-axis shows the -log10(FDR) values of the association results from the previous EWAS meta-analysis of current vs. never smoking*[19]. *The "mQTL-" CpGs were not tested for DNAm → Smoking causal effects in the current study.*

#### References

1. Ligthart L, van Beijsterveldt CEM, Kevenaar ST, de Zeeuw E, van Bergen E, Bruins S, et al. The Netherlands Twin Register: Longitudinal Research Based on Twin and Twin-Family Designs. Twin Research and Human Genetics. 2019;22(6):623–36.

2. Minică CC, Dolan CV, Boomsma DI, De Geus E, Neale MC. Extending Causality Tests with Genetic Instruments: An Integration of Mendelian Randomization with the Classical Twin Design. Behavior Genetics. 2018;48(4):337–49.

3. Castro-de-Araujo LFS, Singh M, Zhou Y, Vinh P, Verhulst B, Dolan CV, et al. MR-DoC2: Bidirectional Causal Modeling with Instrumental Variables and Data from Relatives. Behavior Genetics. 2023 Feb 1;53(1):63–73.

4. Singh M, Verhulst B, Vinh P, Zhou Y (Daniel), Castro-de-Araujo LFS, Hottenga JJ, et al. Using Instrumental Variables to Measure Causation over Time in Cross-Lagged Panel Models. Multivariate Behavioral Research. 2024 Feb 15;59(2):342–70.

5. Auton A, Abecasis GR, Altshuler DM, Durbin RM, Abecasis GR, Bentley DR, et al. A global reference for human genetic variation. Nature. 2015 Oct 1;526(7571):68–74.

6. Haplotype Reference Consortium. A reference panel of 64,976 haplotypes for genotype imputation. Nature Genetics. 2016 Oct 1;48(10):1279–83.

7. Francioli LC, Menelaou A, Pulit SL, van Dijk F, Palamara PF, Elbers CC, et al. Whole-genome sequence variation, population structure and demographic history of the Dutch population. Nature Genetics. 2014 Aug 1;46(8):818–25.

8. Price AL, Patterson NJ, Plenge RM, Weinblatt ME, Shadick NA, Reich D. Principal components analysis corrects for stratification in genome-wide association studies. Nature Genetics. 2006 Aug 1;38(8):904–9.

9. Bibikova M, Barnes B, Tsan C, Ho V, Klotzle B, Le JM, et al. High density DNA methylation array with single CpG site resolution. Genomics. 2011 Oct 1;98(4):288–95.

10. van Dongen J, Nivard MG, Willemsen G, Hottenga JJ, Helmer Q, Dolan CV, et al. Genetic and environmental influences interact with age and sex in shaping the human methylome. Nature Communications. 2016 Sep 1;7(1):11115.

11. van Iterson M, Tobi EW, Slieker RC, den Hollander W, Luijk R, Slagboom PE, et al. MethylAid: visual and interactive quality control of large Illumina 450k datasets. Bioinformatics. 2014 Dec 1;30(23):3435–7.

12. Sinke L, van Iterson M, Cats D, Slieker R, Heijmans B. DNAmArray: Streamlined workflow for the quality control, normalization, and analysis of Illumina methylation array data [Internet]. Zenodo; 2019. Available from: https://doi.org/10.5281/zenodo.3355292

13. Fortin JP, Labbe A, Lemire M, Zanke BW, Hudson TJ, Fertig EJ, et al. Functional normalization of 450k methylation array data improves replication in large cancer studies. Genome Biology. 2014 Dec 3;15(11):503.

14. van Dongen J, Bonder MJ, Dekkers KF, Nivard MG, van Iterson M, Willemsen G, et al. DNA methylation signatures of educational attainment. npj Science of Learning. 2018 Mar 23;3(1):7.

15. Min JL, Hemani G, Hannon E, Dekkers KF, Castillo-Fernandez J, Luijk R, et al. Genomic and phenotypic insights from an atlas of genetic effects on DNA methylation. Nature Genetics. 2021 Sep 1;53(9):1311–21.

16. Chang CC, Chow CC, Tellier LC, Vattikuti S, Purcell SM, Lee JJ. Second-generation PLINK: rising to the challenge of larger and richer datasets. GigaScience. 2015 Dec 1;4(1).

17. Saunders GRB, Wang X, Chen F, Jang SK, Liu M, Wang C, et al. Genetic diversity fuels gene discovery for tobacco and alcohol use. Nature. 2022 Dec 22;612(7941):720–4.

18. Vilhjálmsson J, Yang J, Finucane K, Gusev A, Lindström S, Ripke S, et al. Modeling Linkage Disequilibrium Increases Accuracy of Polygenic Risk Scores. The American Journal of Human Genetics. 2015 Oct 1;97(4):576–92.

19. Joehanes R, Just AC, Marioni R, Pilling L, Reynolds L, Mandaviya PR, et al. Epigenetic Signatures of Cigarette Smoking. Circulation Cardiovascular genetics. 2016;9(5):436–47.

20. Neale MC, Hunter MD, Pritikin JN, Zahery M, Brick TR, Kirkpatrick RM, et al. OpenMx 2.0: Extended Structural Equation and Statistical Modeling. Psychometrika. 2016;81(2):535–49.

21. Verhulst B, Neale MC. Best Practices for Binary and Ordinal Data Analyses. Behavior Genetics. 2021;51(3):204–14.

22. van Iterson M, van Zwet EW, Heijmans BT, the BIOS Consortium. Controlling bias and inflation in epigenome- and transcriptome-wide association studies using the empirical null distribution. Genome Biology. 2017 Jan 27;18(1):19.

23. Gogarten SM, Bhangale T, Conomos MP, Laurie CA, McHugh CP, Painter I, et al. GWASTools: an R/Bioconductor package for quality control and analysis of genome-wide association studies. Bioinformatics. 2012 Dec 1;28(24):3329–31.

24. Benjamini Y, Hochberg Y. Controlling the False Discovery Rate: A Practical and Powerful Approach to Multiple Testing. Journal of the Royal Statistical Society: Series B (Methodological). 1995 Jan 1;57(1):289–300.

25. Storey J, Bass A, Dabney A, Robinson D. qvalue: Q-value estimation for false discovery rate control. doi:10.18129/B9.bioc.qvalue [Internet]. 2023. Available from: https://doi.org/10.18129/B9.bioc.qvalue

26. Zhou Y, Zhou B, Pache L, Chang M, Khodabakhshi AH, Tanaseichuk O, et al. Metascape provides a biologist-oriented resource for the analysis of systems-level datasets. Nature Communications. 2019 Apr 3;10(1).

27. Ashburner M, Ball CA, Blake JA, Botstein D, Butler H, Cherry JM, et al. Gene Ontology: tool for the unification of biology. Nature Genetics. 2000 May 1;25(1):25–9.

28. Kanehisa M, Goto S. KEGG: Kyoto Encyclopedia of Genes and Genomes. Nucleic Acids Research. 2000 Jan 1;28(1):27–30.

29. Subramanian A, Tamayo P, Mootha VK, Mukherjee S, Ebert BL, Gillette MA, et al. Gene set enrichment analysis: A knowledge-based approach for interpreting genome-wide expression profiles. Proceedings of the National Academy of Sciences. 2005 Oct 25;102(43):15545–50.

30. Fabregat A, Jupe S, Matthews L, Sidiropoulos K, Gillespie M, Garapati P, et al. The Reactome Pathway Knowledgebase. Nucleic Acids Research. 2018 Jan 4;46(D1):D649–55.

31. Breeze CE, Paul DS, van Dongen J, Butcher LM, Ambrose JC, Barrett JE, et al. eFORGE: A Tool for Identifying Cell Type-Specific Signal in Epigenomic Data. Cell Reports. 2016 Nov 15;17(8):2137–50.

32. Breeze CE, Reynolds AP, van Dongen J, Dunham I, Lazar J, Neph S, et al. eFORGE v2.0: updated analysis of cell type-specific signal in epigenomic data. Bioinformatics. 2019 Nov 15;35(22):4767–9.

33. Breeze CE. Cell Type-Specific Signal Analysis in Epigenome-Wide Association Studies. In: Guan W, editor. Epigenome-Wide Association Studies: Methods and Protocols [Internet]. New York, NY: Springer US; 2022. p. 57–71. Available from: https://doi.org/10.1007/978-1-0716-1994-0_5

34. Vuckovic D, Bao EL, Akbari P, Lareau CA, Mousas A, Jiang T, et al. The Polygenic and Monogenic Basis of Blood Traits and Diseases. Cell. 2020 Sep 3;182(5):1214-1231.e11.
